# Genetic overlap between mood instability and alcohol-related phenotypes suggests shared biological underpinnings

**DOI:** 10.1101/2022.03.08.22271329

**Authors:** Romain Icick, Alexey Shadrin, Børge Holen, Naz Karadag, Aihua Lin, Guy Hindley, Kevin O’Connell, Oleksandr Frei, Shahram Bahrami, Margrethe Collier Høegh, Weiqiu Cheng, Chun C. Fan, Srdjan Djurovic, Anders M. Dale, Trine Vik Lagerberg, Olav B. Smeland, Ole A. Andreassen

**Affiliations:** NORMENT Centre, Institute of Clinical Medicine, University of Oslo and Division of Mental Health and Addiction, Oslo University Hospital, 0407 Oslo, Norway; Université de Paris, INSERM UMR-S1144, F-75006, France; Center for Bioinformatics, Department of Informatics, University of Oslo, PO box 1080, Blindern, 0316 Oslo, Norway; Department of Radiology, University of California, San Diego, La Jolla, CA 92093, United States of America; Multimodal Imaging Laboratory, University of California San Diego, La Jolla, CA 92093, United States of America; Department of Cognitive Science, University of California, San Diego, La Jolla, CA, USA; Department of Medical Genetics, Oslo University Hospital, Oslo, Norway; NORMENT Centre, Department of Clinical Science, University of Bergen, Bergen, Norway; KG Jebsen Centre for Neurodevelopmental disorders, University of Oslo, Oslo, Norway; Department of Psychiatry, University of California, San Diego, La Jolla, CA, USA; Department of Neurosciences, University of California San Diego, La Jolla, CA 92093, United States of America

**Author notes:** **Corresponding author:** Romain Icick, NORMENT Centre, building 49, Oslo University Hospital, Ullevål, Kirkeveien 166, 0424 Oslo, Norway.

**Keywords:** Polygenic overlap, joint genome-wide association study, mood instability, alcohol consumption, alcohol use disorder

## Abstract

Alcohol use disorder (AUD) is a pervasive and devastating mental illness with high comorbidity rates with other mental disorders. We aimed to characterize the shared *vs*. unique polygenicity of AUD, alcohol consumption (AC) and mood instability (MOOD), a relevant transdiagnostic factor, using large genome-wide association studies (GWASs) data. We hypothesize that cross-analyzing these phenotypes would shed light on their unique and shared polygenicity, increase our knowledge regarding the genetic basis of the comorbidity between AUD and mood disorders, and boost discovery for jointly-associated loci. Summary statistics for MOOD, AC and AUD GWASs (Ns =363,705; 200,680 and 200,004; respectively) were analysed to characterize the cross-phenotype associations between MOOD and AC, MOOD and AUD and AC and AUD, respectively. To do so, we used a newly-established pipeline that combines (i) the bivariate causal mixture model (MiXeR) to quantify the cross-phenotype polygenic overlap and (ii) the conjunctional false discovery rate (conjFDR) to discover specific jointly-associated genomic loci. These loci were functionally characterized and mapped to genes and biological functions. We also performed validation in independent samples and phenotypic analyses. MOOD was highly polygenic (10,400 single nucleotide polymorphisms, SNPs) compared to AC and AUD (4,900 SNPs, SD =600 and 4,300 SNPs, SD =2,000; respectively). The polygenic overlap of MOOD and AC was much larger than that of MOOD and AUD (98% *vs*. 49%) and genetic correlation was opposite (−0.2 *vs*. 0.23), which was confirmed in independent samples. MOOD&AUD causal SNPs were significantly enriched for brain genes, conversely to MOOD&AC. Among 38 loci identified in the joint analysis, sixteen were novel for MOOD, AC and AUD. Similarly distinct patterns were evidenced for SNP localization, function and previous GWAS associations outside of the phenotypes that were currently studied. MOOD, AC and AUD were also strongly associated at the phenotypic level. Overall, using multilevel polygenic quantification, joint loci discovery and functional annotation methods, we evidenced that the polygenic overlap between MOOD and AC/AUD implicated shared biological underpinnings but clearly distinct functional patterns between MOOD&AC and MOOD&AUD. Using the MOOD endophenotype, the current study suggests new mechanisms for the comorbidity of AUD with mood disorders.

## INTRODUCTION

Alcohol use disorder (AUD) is a mental disorder characterized by a chronic loss of control over the use of alcohol. Alcohol is the most burdensome of addictive substances, contributing to three million deaths each year (Global Burden of Disease, World Health Organization 2018). Alcohol consumption (AC) and AUD do not share the same clinical phenomenology [1], neuropsychology [2], or biological underpinnings - whether on a neuroimaging [3] or a genetic basis [4–6]. AUD is diagnosed in 15% of regular alcohol users [7], suggesting that AC is a necessary, yet far from sufficient behavior for causing AUD [8]. Despite these differences, however, AC and AUD show substantial genetic correlation ∼0.5 [8]. Thus, taken altogether, these data encourage further investigations of the shared and unique molecular underpinnings of alcohol consumption (AC) *vs*. AUD using genetic analyses.

Accumulating evidence for high genetic correlation [4,9] and shared environmental factors between AUD and non-substance-related psychiatric disorders [10] have placed greater emphasis on the transdiagnostic vulnerability to mental disorders. Mood instability (MOOD), both a risk factor and a clinical expression of psychiatric disorders [11,12], represents one of these transdiagnostic factors. MOOD has been associated with AUD at the clinical and at the genetic level [13–15], and is a common feature of the psychiatric comorbidities that are commonly associated with AUD, including mood [16], psychotic [17], anxiety [18], and personality [19] disorders. Importantly for the current study, it has further been associated with increased MOOD in a sample of patients with bipolar disorder [20]. Likewise, clinical studies have shown a strong bi-directional relationship between MOOD and AC [21–24]. Therefore, the interaction of MOOD and AC likely increases the risk of developing mood disorders and/or AUD [25]. With that regard, studying the genetic liability of MOOD and AC *vs*. MOOD and AUD promises to yield new insights into the pathophysiology of AUD and open avenues to better model the comorbidity between mood disorders and AUD. These mechanistic hypotheses fit particularly well with the research domain criteria initiative [RDoC [26]]. Under this framework, MOOD could represent a transdiagnostic factor (arousal construct), which might facilitate the transition from regular AC to AUD by increasing the liability toward maladaptive habits (reward learning construct), especially in the context of premorbid impairments in the reward responsiveness construct. Eventually, characterizing the unique *vs*. shared polygenic liability of these traits and disorder is thus of paramount importance, yet remains unclear [27].

Significant genetic correlation has been shown between mood disorders and AUD [8], between MOOD and mood disorders [28], but not between AC and mood disorders [8]. However, genetic correlation measures only provide a summary measure of genome-wide correlation of effect sizes. They have therefore been unable to quantify the extent of shared versus unique heritability between traits and disorders, the extent of shared genetic factors with concordant and discordant effects on each pair of traits, or identify specific jointly-associated genomic loci. Further triangulating these traits and disorders may unravel specific relationships across core mental features, especially by extending the search for shared vulnerability between psychiatric phenotypes [29] - beyond genetic correlation. Building on prior findings about genetic correlations between MOOD and AUD, our group applied the bivariate causal mixture model (MiXeR) to quantify the amount of genetic overlap [30], and conjunctional FDR (conjFDR) to identify specific overlapping vulnerability loci [31] to relevant largest-to-date GWASs. For instance, conjFDR has been successfully applied to AC *vs*. AUD and bipolar disorder or schizophrenia, showing interesting mixed effect directions [32] and calling for extension using transdiagnostic dimensions such as MOOD.

Our aim was to determine the unique *vs*. shared polygenic liability between AC, an alcohol-related trait, and AUD, an alcohol-related disorder. To do so, we characterized their overlapping and unique polygenic liability with a transdiagnostic trait, MOOD. We hypothesized that triangulating these traits and disorder beyond genetic correlation would address issues regarding biological pathways relevant to AUD as compared to AC. We also hypothesized that having MOOD as a primary trandisagnostic phenotype would open avenues toward a better understanding of the comorbidity between mood disorders and AUD.

## MATERIAL AND METHODS

We applied a newly-established pipeline for characterizing the polygenic architecture of MOOD and AC or AUD, based on cross-trait overlap and improved power for the discovery of overlapping loci using. We used the bivariate causal mixture model (MiXeR), and the conjunctional FDR method (conjFDR), investigating genome-wide association studies (GWAS) summary statistics data from relevant phenotypes. The manuscript follows the statement of STrengthening the REporting of Genetic Association studies (STREGA) [33]. Effective sample sizes and SNPs are summarized in **Table 1**. More methodological details are given in the **Supplementary Methods File**.

**Table 1:**
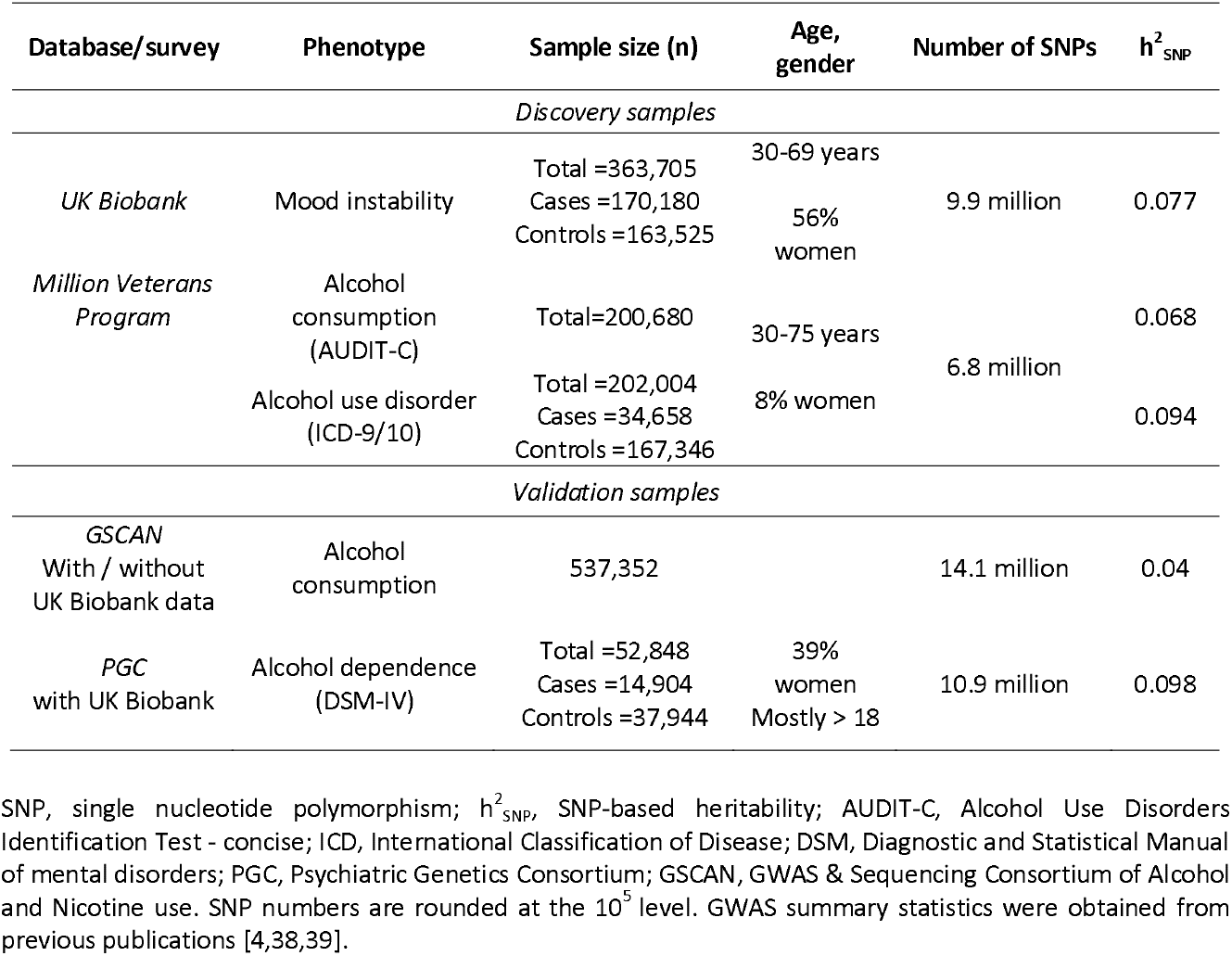
Characteristics of original GWASs on mood instability and alcohol-related phenotypes.

| Database/survey | Phenotype | Sample size (n) | Age, gender | Number of SNPs | h<sup>2</sup><sub>SNP</sub> |
| --- | --- | --- | --- | --- | --- |
| Discovery samples |  |  |  |  |  |
| UK Biobank | Mood instability | Total =363,705<br>Cases =170,180<br>Controls =163,525 | 30-69 years<br>56% women | 9.9 million | 0.077 |
| Million Veterans Program | Alcohol consumption (AUDIT-C) | Total=200,680 | 30-75 years | 6.8 million | 0.068 |
|  | Alcohol use disorder (ICD-9/10) | Total =202,004<br>Cases =34,658<br>Controls =167,346 | 8% women |  | 0.094 |
| Validation samples |  |  |  |  |  |
| GSCAN<br>With / without UK Biobank data | Alcohol consumption | 537,352 |  | 14.1 million | 0.04 |
| PGC<br>with UK Biobank | Alcohol dependence (DSM-IV) | Total =52,848<br>Cases =14,904<br>Controls =37,944 | 39% women<br>Mostly > 18 | 10.9 million | 0.098 |

SNP, single nucleotide polymorphism; $h^2_{\text{SNP}}$ , SNP-based heritability; AUDIT-C, Alcohol Use Disorders Identification Test - concise; ICD, International Classification of Disease; DSM, Diagnostic and Statistical Manual of mental disorders; PGC, Psychiatric Genetics Consortium; GSCAN, GWAS & Sequencing Consortium of Alcohol and Nicotine use. SNP numbers are rounded at the $10^5$ level. GWAS summary statistics were obtained from previous publications [4,38,39].

### Samples and phenotypes

We extracted effect sizes and *p*-values from GWASs performed on each of the selected phenotypes. All participants were of European ancestry in order to maintain linkage disequilibrium (LD) homogeneity across samples, since inconsistency of LD between the reference panel and GWAS sample may bias both MiXeR and conjFDR outcomes. Briefly, the genotyped sample of the UK Biobank consists of 488,377 subjects from the community assessed for a wide range of measures. The Million Veterans Program (MVP) consists of a dataset of more than 200,000 genotyped individuals from the US Veterans Health Care System, whose phenotypic information was obtained during clinical appointments and was extensively recorded in electronic health records. The recruitment and genetic analysis for both samples required written informed consent. They fulfilled ethical standards of the Helsinki declaration 1989. Recruitment was approved by the National Health Service National Research Ethics Service, Ref. 16/NW/0274, and genetic analyses by the Veteran Affairs Central Institutional Review Board.

The primary phenotype was MOOD, defined in the UK Biobank by answering yes/no to the question “does your mood often go up and down?”, as used in a previously-published GWAS [28]. Despite the simplicity of this measure, it was evidenced to be the strongest predictive factor for both bipolar and major depressive disorder in a prospective community sample [12]. More generally, single-item measurements of sadness and psychological distress have been found to be useful in the general population [34] and in clinical samples with recurrent depression [35]. Finally, the reliable identification of cross-trait genetic overlap requires very large sample sizes that were not available with more extensive measurements of mood/affective instability at the time of the current study [36].

We focused on two secondary alcohol-related phenotypes: a continuous trait, AC, and a binary disorder, AUD from the MVP survey [37]. AC was measured using the Alcohol Use Disorders Identification Test-Concise (AUDIT-C), which measures typical quantity (item 1) and frequency (item 2) of drinking, and frequency of heavy or binge drinking (item 3) over the past year. AUDIT-C yields a continuous score ranging from 0 to 12. AUD was defined as having at least one inpatient or two outpatient diagnoses of alcohol abuse or dependence or two diagnoses of alcohol intoxication according to the International Classification of Disease (ICD)-9 or -10.

### Statistical analysis

Firstly, we used MiXeR to quantify total polygenic overlap between MOOD and alcohol-related phenotypes (AC, AUD) at the genome-wide level [30]. The method takes GWAS summary statistics on two phenotypes and provides maximum likelihood estimates for the total number of shared and phenotype-specific for non-null variants, and genetic correlation between phenotypes accounting for sample overlap. Secondly, we estimated cross-phenotype enrichment using conditional Q-Q plots [31]. Conditional Q-Q plots are a model-free approach for visual assessment of shared genetic background between two phenotypes which provides complementary support for MiXeR estimates of genetic overlap. Thirdly, we performed conjunctional false discovery rate analysis (conjFDR) [31] to discover specific genomic loci jointly associated with each alcohol-related phenotype (AC, AUD) and MOOD. In the specific context of the current study, conjFDR was deemed particularly relevant in order to identify genomic loci associated with MOOD that were either shared or unique to each of the alcohol-related phenotypes.

Statistical analyses were conducted on Linux^®^-based servers. Plotting and downstream analyses were performed with R 3.6.3 and R Studio 1.4.1103.

### Genomic loci definition

We defined genomic loci according to the Functional Mapping and Annotation of Genome-Wide Association Studies (FUMA GWAS, https://fuma.ctglab.nl/), based on the r^2^ statistic for measuring LD.

### Functional annotation

Candidate SNPs for each set of conjFDR analysis (*p* <0.1) were annotated with FUMA in three steps:

i. Mapping loci to SNPs and SNPs to genes based on positional, gene expression and chromatin interaction data (FUMA *SNP2GENE*) - yielding a list hereafter referred to as *credibly mapped genes*;
ii. Annotating each candidate SNP for:
  a. Combined Annotation Dependent Depletion (CADD) scores, which predict SNP dleteriousness on protein structure/function;
  b. RegulomeDB scores, which predict regulatory functionality;
  c. Chromatin states, which predict transcription/regulatory effects from chromatin states at the SNP locus;
  d. Novelty, based on candidate genomic ranges compared to the latest build of GWAScatalog + MEDLINE search.
iii. Passing the resulting list of genes into FUMA *GENE2FUNCTION* to estimate gene-set enrichment. All analyses were corrected for multiple comparisons using the Benjamini-Hochberg method for each pathway/category. For the sake of brevity, we chose to restrict the enrichment testing of credibly mapped genes for positional gene sets, canonical pathways and gene expression as a function of tissue (GTEx data v.8 https://gtexportal.org/home/) and developmental age (https://www.brainspan.org/). Tissue and age-specific gene expression are considered premium datasets in order to describe the functional genomics associated with GWAS data, thus providing valuable insights into psychiatric genomics (see [40] for review);
iv. Celltype specificity using SNP-epigenomics associations from the *Roadmap* (http://www.roadmapepigenomics.org/) and the *ENCODE* (https://www.encodeproject.org/) databases;
v. DrugBank database, providing hints regarding the druggability of credibly mapped genes and clues for possible drug repurposing.

An overview of the statistical pipeline is provided in **Supplementary Figure 1**, and further details are provided in the following references [31,41].

### Validation in independent alcohol GWASs

We reanalyzed the MOOD x AC and MOOD x AUD relationships using the largest and most recent independent GWASs summary statistics for alcohol-related phenotypes. This included AUD from the Psychiatric Genetics Consortium (AUD-PGC) and AC from the GWAS & Sequencing Consortium of Alcohol and Nicotine use (AC-GSCAN). We also tested the concordance of the effect directions for each lead SNP between the discovery analyses and the re-analyses, using exact binomial tests. The test yields the probability to get observed number of concordant SNPs assuming effect directions are selected randomly.

### Associations between MOOD, AC and AUD at the phenotypic level

To complete the characterization of the polygenic overlap between our phenotypes of interest, we leveraged the UK Biobank phenotypic data (accession number 27412) to analyse associations between MOOD, AC and AUD, using a series of regression models adjusted for sex, age, and the 20 first genetic principal components.

## RESULTS

A complete overview of the shared polygenicity and genetic correlation for each pair of phenotypes is shown in **Supplementary Table 2**. Results from the validation analysis are available as supplementary data (text, **Supplementary Table 3** and **Supplementary Figures 7-11**).

### Trait-specific and shared genetic architecture (MiXeR)

**Figure 1** illustrates the high polygenicity of MOOD (10.4k SNPs) as compared to the moderate polygenicity of AC (4.9k SNPs) and AUD (4.3k SNPs). MiXeR showed that almost all AC loci overlapped with MOOD liability whereas only ∼50% of AUD genes did so, despite the large overlap in genetic architecture between AC and AUD. This complex picture of mixed effect directions was also in line with the genetic correlation (r_g_) results, which was significant for all pairs of phenotypes: moderately negative (r_g_ = -0.22) for MOOD and AC (**Figure 1A**), moderately positive (r_g_ = 0.23) for MOOD and AUD (**Figure 1A)**, and strongly positive between AC and AUD (r_g_ =0.52, **Figure 1B**). This indicates different directions of effects between the shared polygenic variants of MOOD and AC versus MOOD and AUD and suggests different molecular mechanisms emerging from partly similar pathways. MiXeR results for MOOD and AC were of high confidence revealing clear optimum in log-likelihood plot (**Supplementary Figure 2A**) reflected in tight SD and supported by positive AIC value when compared to the model with minimum overlap. However, the results involving the MVP AUD sample should be interpreted with caution because of erratic behaviour of log-likelihood profiles across 20 iterations of the analysis (**Supplementary Figure 2, B and C**) resulting in high SD of modelled estimates. Importantly, however, in both MOOD and AUD, and AC and AUD analyses, AIC values were positive comparing modelled overlap to complete overlap and negative comparing modelled estimate to minimum possible overlap. Therefore, in both analyses, AIC supports the existence of AUD-specific fraction of “causal” variants.

**Figure 1:**
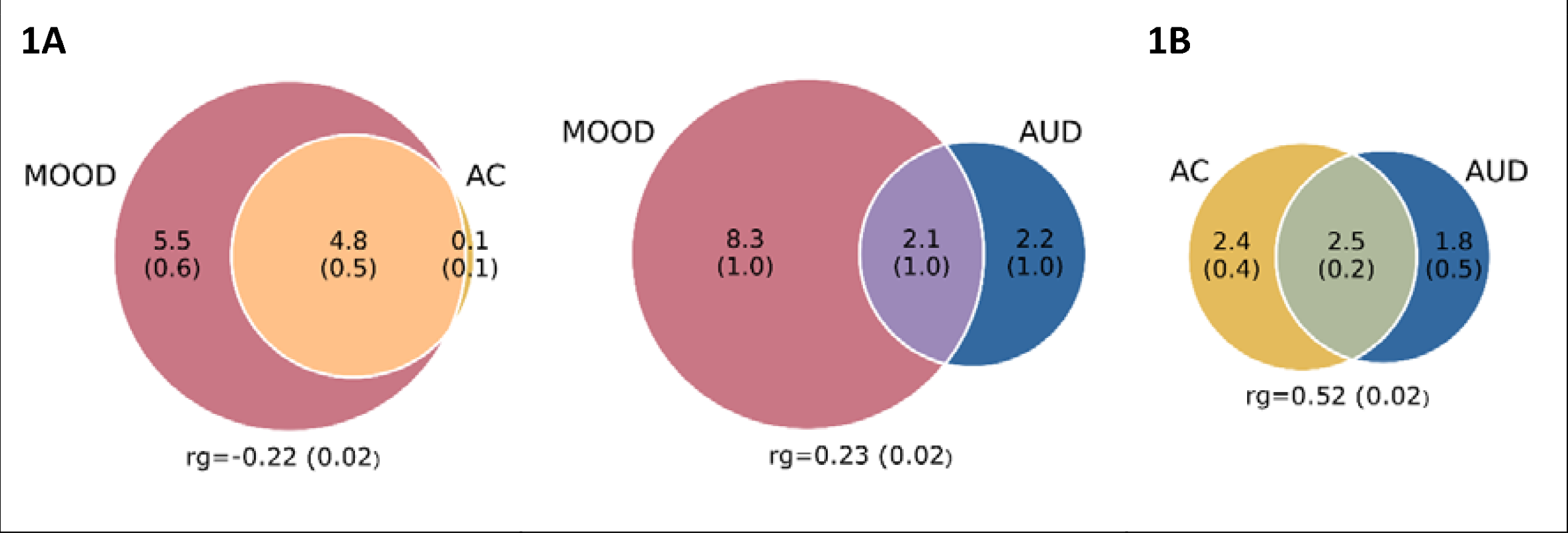
overlapping ‘causal’ SNPs (in thousands) explaining 90% of heritability and genetic correlation with corresponding standard deviations in parenthesis between (A) mood instability (MOOD) and alcohol consumption (AC) and MOOD and alcohol use disorder (AUD) and (B) between AC and AUD (B). MOOD, mood instability in the UK Biobank; AC, alcohol consumption in the MVP sample; AUD, alcohol use disorder in the MVP sample; rg, genetic correlation.

Likewise, there was significant enrichment of SNPs associated with MOOD when conditioning on AC and AUD and vice-versa, as evidenced by increasing deviation from the expected null line of SNP strata with increasing significance in the conditional trait on Q-Q plots (**Supplementary Figures 3A&3B**).

### Loci discovery and annotation

#### Loci discovery (conjFDR)

At conjFDR *p*<0.05, there were 18 significant lead SNPs in the AC&MOOD analysis and 20 in the AUD&MOOD analysis. Manhattan plots for conjFDR of MOOD&AC and MOOD&AUD analyses are shown in **Figure 2**. Among the 38 lead SNPs, only one (rs2312147) was common to both AC and AUD. Functional annotation of these genome-wide significant SNPs is available on **Supplementary Table 1**.

**Figure 2:**
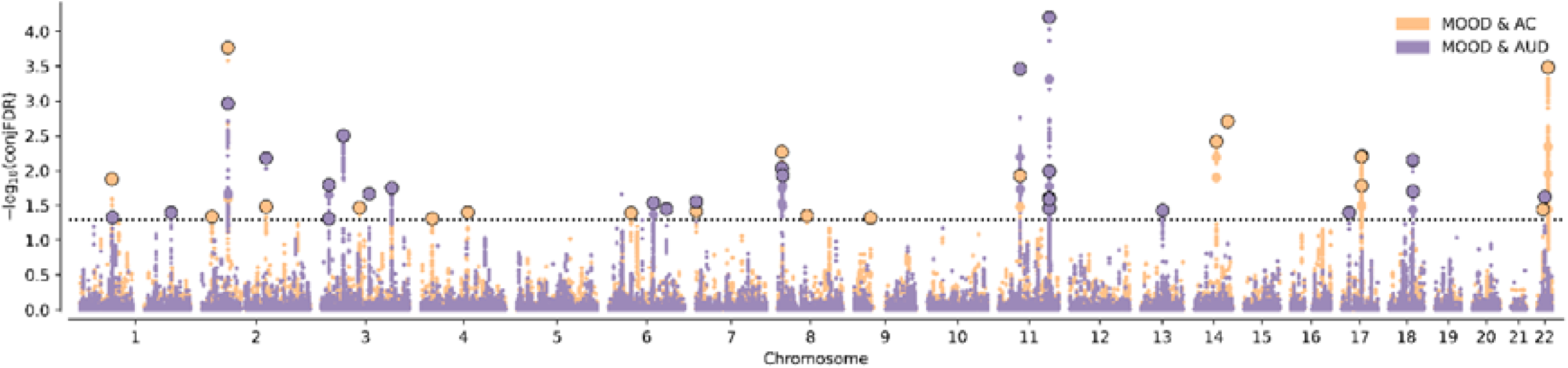
combined conjFDR Manhattan plot for MOOD&AC and MOOD&AUD. FDR, false discovery rate; MOOD, mood instability in the UK Biobank; AC, alcohol consumption in the MVP sample; AUD, alcohol use disorder in the MVP sample.

Sixteen SNPs were novel with regards to MOOD, AC and AUD GWASs that had been published as of January 28, 2022. Three of these SNPs had potential functional impact due to: deleteriousness (rs34811474, CADD score =23.7), non-synonymous protein effect (rs8007859) and location in a regulatory region (UTR3, rs11130187). None of the novel SNPs were shared between AC and AUD. Interestingly, one of these SNPs (rs2277840, MOOD&AUD conjFDR) had been associated with the level of response to alcohol, an endophenotype for AUD, in a small-scale GWAS [42].

**Table 2:**
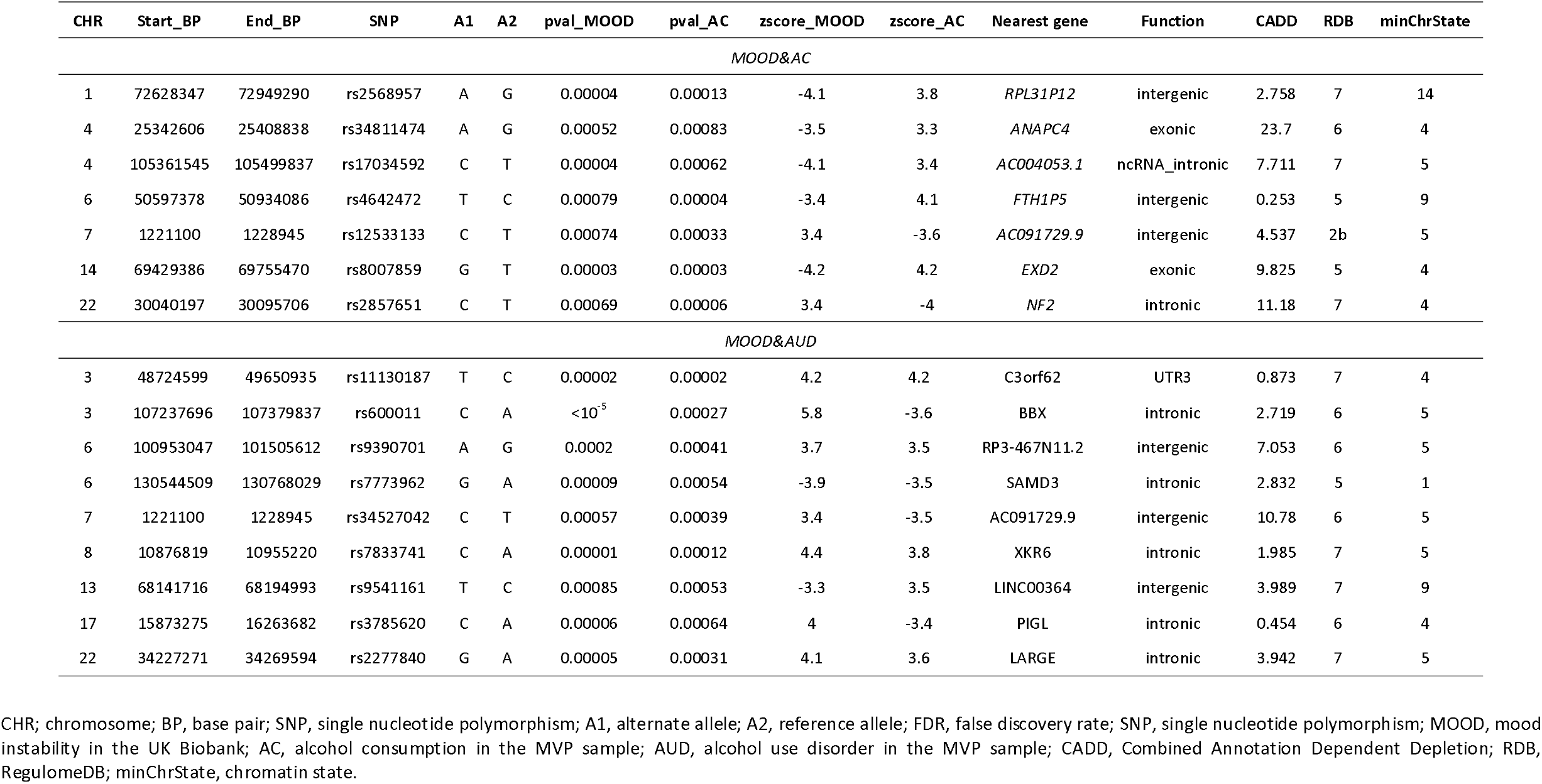
Significantly associated SNPs for MOOD&AC and for MOOD&AUD, by conjFDR, previously unpublished in GWASs of MOOD, AC or AUD.

#### Functional analyses of lead SNPs by FUMA

The pattern of genomic localization of SNPs associated with MOOD and alcohol-related phenotypes differed between AC and AUD. In the MOOD&AC analysis, SNPs were significantly enriched for 3’ and 5’ untranslated regions (*p* =3 × 10^−4^ and 9 × 10^−8^ , respectively) and downstream areas (*p* =0.002) while SNPs from the MOOD&AUD analysis were significantly enriched for intronic (*p* =10^−315^), intergenic (*p* =10^−152^), exonic coding regions (*p* =0.04), non-coding long RNA (*p* =0.009), and upstream regions (*p* =0.009).

According to FUMA the function of the credibly mapped genes from conjFDR mostly differed for MOOD&AC *vs* MOOD&AUD - in line with MiXeR overall patterns. Firstly, genes from the MOOD&AC analysis shared enrichment only for the 11p11 region. Secondly, the *reactome* post translational protein modification pathway was significantly enriched for both alcohol-related traits, but the MOOD&AC analysis elicited two more pathways related to the TFAP2 family of transcription factors (**Supplementary Figure 4**). Thirdly, in the MOOD&AC analysis, the hypothalamus was the only brain tissue with significant enrichment in gene expression, whereas, in the MOOD&AUD analysis, all but four brain tissues were significantly overrepresented (**Supplementary Figures 5A&5B**) with downregulated genes. Fourthly, for AC (**Supplementary Figure 6A**), only 12 genes (18%) showed a progressively decreasing brain expression throughout the lifespan while 19 (28%) showed increasing expression. For AUD, eight (16%) genes show increasing expression throughout the lifespan while 26 (33%) show progressively reduced expression (Chi^2^ *p* =0.031, **Supplementary Figures 6A&6B**). There was no difference between conjFDR analyses in celltype enrichment using epigenetics databases. Although their number was too low to perform a complete functional analysis of the 16 novel SNPs, passing them through GWAScatalog showed that cognitive traits were only represented in MOOD&AC, while affective traits were only represented in MOOD&AUD, durther supporting the distinct neurobiological patterns described above (**Supplementary Table 4**). Finally, nine genes from the MOOD&AC analysis and 12 from the MOOD&AUD yielded hits in *DrugBank*. Among the latter, there were multiple *DrugBank* hits for the genes encoding Inosine monophosphate dehydrogenase 2 (*IMPDH2*, eight hits), the type 2 dopamine receptor (*DRD2*, 110 hits) - a classical target in mental health, and the type 2 adenosine receptor (*ADORA2B*, five hits), one of the targets of the approved AUD medication acamprosate. Other drugs (one hits per gene) were related to the immune and metabolic systems medications as well as to nutrients. None of the novel associations was included in these hits.

#### Phenotypic associations between MOOD, AC and AUD

After adjustment on sex, age and ancestry, each linear relationship between MOOD and both alcohol-related phenotypes remained strong and significant (all *p* <0.001): AC ∼ MOOD (β =0.003), AUD ∼ MOOD (β =0.62), MOOD ∼ AC (β =0.89) and MOOD ∼ AUD (β =0.62). In the model where MOOD and AC were entered together as independent variables, AUD was significantly associated with MOOD (β =0.7), with AC (β =0.03), and with their interaction term (β =-0.005); all *p* <0.001.

## DISCUSSION

We leveraged data from large scale GWASs to shed light on the shared and unique polygenic architecture of MOOD, AC and AUD, suggesting shared biological underpinnings betwee MOOD and alcohol-related phenotypes. We obtained remarkably consistent and converging evidence that the relationship between MOOD and alcohol-related phenotypes strongly differed for AC *vs*. AUD, thus confirming our main hypothesis. Using MiXeR, we first evidenced a large degree of polygenic overlap between MOOD and alcohol-related phenotypes, which was higher for AC (98%) than for AUD (49%). Next, using conjFDR, 20 SNPs for MOOD&AC and 18 for MOOD&AUD reached genome-wide significance. Importantly, none was common to both analyses, and sixteen were novel at the time of the study for any of the phenotypes - based on a highly conservative checking procedure including the latest conjFDR results from our group [32]. At all levels, the functional annotation of conjFDR supported the differences in polygenic overlap between the joint genetic architecture of the vulnerability to MOOD&AC *vs*. MOOD&AUD. Taken altogether, these findings describe a polygenic scenario with distinctive patterns of effect distributions among the overlapping variants, which may help to define the unique features of each phenotype. Our study was performed using the latest GWASs available for two traits of major relevance for mental health, *i*.*e*. MOOD and AC; and characterized how these relate to AUD. The growing number of adequately powered GWASs for alcohol-related phenotypes allowed us to perform a series of validation analyses, which overall strengthened our discovery results as regards polygenic overlap, shared loci identification, and concordance of effect direction.

Our first main finding was that AC and AUD, beyond confirming their strong, positive genetic correlation [32], showed relevant differences in the proportion of shared *vs*. unique polygenicity with MOOD. Strikingly, genetic correlation showed opposite directions (negative for AC, positive for AUD). Unfortunately, available AUD samples (MVP, PGC - and their meta-analysis) did not enable enough stability of MiXeR analysis to further discuss the number of overlapping causal variants with MOOD in detail. However, given the high statistical power achieved by meta-analyzing the MVP+PGC samples, this was unlikely due to power issues. Instead, one could hypothesize that the polygenicity of AUD follows a multimodal distribution (including consistent evidence for peak and pleiotropic signals [4,8,37,43]) which, combined with its low SNP-based heritability (<10%), did not allow to generate clear-cut MiXeR outcomes. Future studies using, e.g., multivariate approaches [44] could help strengthening these findings.

The different polygenic overlap between MOOD&AC *vs*. MOOD&AUD extends genetic correlation results [38] showing that AUDIT-P (a proxy for AUD), but not AC, was associated with most other psychiatric disorders. Additionally, the current findings extend a recent report regarding the polygenicity of AC and AUD and their relationship with schizophrenia and bipolar disorder [32]. Four (out of 38) of the loci that we identified were also reported in this previous study (out of 62). Firstly, this suggests a consistent power for locus discovery between the two studies, that is, 15-20 loci per pair of phenotypes. Secondly, we notice that eight loci identified as novel in the current study were not reported by Wiström et al. [32] and that this previous study did not identify different biological pathways nor gene expression patterns when using either AC or AUD as a secondary phenotype, despite identifying a higher number of loci. Overall, this supports both the reliability of conjFDR, but also its sensitivity to the phenotypes that are investigated. With that regard, we notice that all of the SNPs we report as novel would reach Bonferroni-corrected genome-wide significance (highest *p*-value =4.8 × 10^−8^), suggesting a true boost in discovery power of conjFDR instead of an effect of relaxing the initial significance threshold for candidate loci. The relevance of combining endophenotypes such as MOOD with disorders such as AUD, which has already been suggested by our group [45], should thus be further explored using RDoC constructs.

Strikingly, the current genetic findings support the phenomenological differences between alcohol use (AC) and alcohol use disorders (AUD). AC (even when regular and in relatively high amounts) is a normative behavior in most Western world countries, while AUD can develop into a pervasive, devastating mental illness. Half of the polygenic overlap and only two loci and none of the functional analysis results were shared between MOOD&AC and MOOD&AUD conjFDR in our study (although this was based on 38 loci in total as compared to 5-10k representing the overall polygenic signal). Thus, the loci associated with MOOD&AUD were significantly enriched for genes that are typically under-expressed in the putamen, amygdala, substantia nigra and anterior cingulate cortex, which was not the case for any of the MOOD&AC loci. Strikingly, these brain regions are at the core of the neurocircuitry of AUD, bot not that of AC [3,46–48]. Additionally, the genes associated with MOOD&AC showed mostly increasing expression throughout brain development, while a significantly opposite trend was observed for the genes associated with MOOD&AUD. These findings are of paramount importance with regard to the underlying neurobiology of AC compared to that of AUD. There is now consistent evidence that AUD develops from AC through the progressive recruitment of extended brain networks [49] implying different neurotransmitter systems [50].

The significant negative correlation found between MOOD and AC was somehow unexpected. At the phenotypic level, both AC and AUD have been strongly associated with MOOD, including in the current original analysis. In clinical samples, the association of MOOD and AC was bi-directional and involved both negative and positive reinforcement [51,52]. Here, we tentatively suggest that individuals with a high genetic load for MOOD will exhibit high levels of AC due to the resulting MOOD phenotype, regardless of their genetic liability toward AC. Accordingly, individuals with a high genetic load for AC may drink more alcohol, whatever their level of genetic vulnerability to MOOD.

Our study has limitations. Firstly, both the MOOD and AC traits remain relatively non-specific and may be heterogeneous, although they represent highly relevant constructs for research. We hope that large GWASs will be conducted based on finer-grained assessment of MOOD in the future. Secondly, the MVP sample is composed of 92% men aged >30, all with military duty, and we could not obtain the phenotypic breakdown for this sample. This limits the generalizability of our findings, since both the prevalence and severity of MOOD, AC and AUD are susceptible to change over the lifespan [14,15,53]. In line with this, we only considered European ancestry subsamples for analysis. We plan to develop trans-ancestral overlap analyses in the near future to overcome this important limitation for better precision psychiatry. Thirdly, the results regarding AUD in the primary analysis should be interpreted with caution given statistical issues (reflected in high SD value for the number of SNPs evidenced by MiXeR), while noticing that additional analyses suggested that power was not involved. Fourthly, MiXeR does not allow for mapping of SNPs involved in the observed polygenic overlap and only takes two traits as input. To overcome this limitation and better describe complex overlap such as reported in the current paper, a trivariate version of MiXeR is currently under development.

## CONCLUSION

By leveraging large GWAS data beyond genetic correlations using MiXeR and conjFDR, the characterization of the polygenic overlap between mood instability and alcohol-related phenotypes improved our understanding of their shared *vs*. unique polygenicity. Our findings open avenues toward a better understanding of the comorbidity between mental disorders, especially between AUD and mood disorders through MOOD, and extend the growing body of evidence regarding how AC and AUD differ in terms of biological mechanisms. The genetic susceptibility to mood instability could represent a missing link between AC and AUD, providing strong biological support to clinical observations.

## Data Availability

Data used in the present study are not publicly available due to restricted access of some datasets.

## AUTHOR CONTRIBUTIONS (CREDIT ROLES)

RI, GH, AS, OF and OAA, Conceptualization; AS, Formal analysis; KOC, SD, AMD, TVL, OS and OAA, Funding acquisition; Methodology, AL, OF, SB, WC, CF, AMD, OS, OAA; NK, Project administration; MCH, SD, TVL, Resources; RI, Writing - original draft; AS, BH, GH, OF, OS, OAA, Writing - review & editing. All authors have critically reviewed and approved the manuscript.

## ACKNOWLEDGEMENTS

The authors would like to thank all the support personnel from the Norwegian center of excellence for mental disorders (NORMENT) and all the researchers that provided valuable input for the current study. The authors would also like the INTPART program for supporting a post-doc exchange for RI.

---

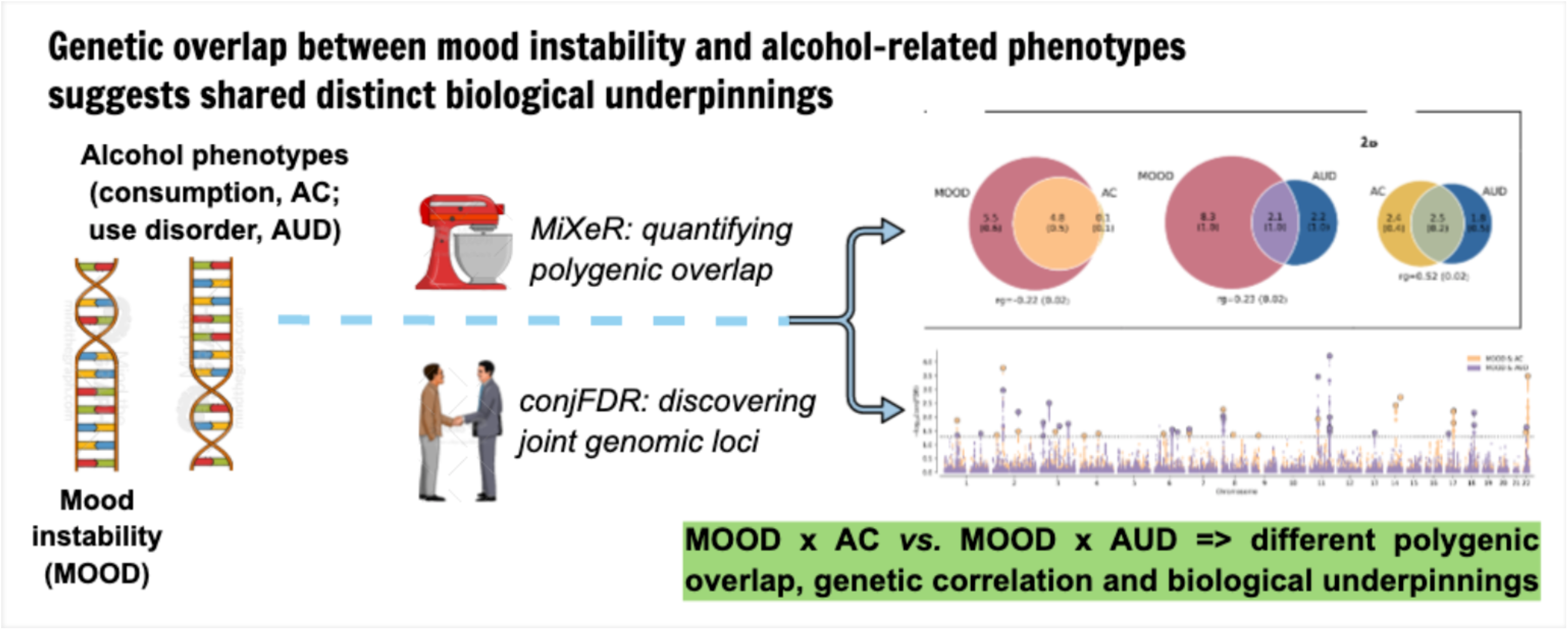

