## Supplementary Methods for "Genetic overlap between mood instability and alcohol-related phenotypes suggests shared biological underpinnings"

Romain Icick et al.

**Supplementary Methods Data**

### MiXeR details

MiXeR assumes that each phenotype has a fraction of non-null ("causal") variants uniformly distributed throughout the genome with effect sizes drawn from the same normal distribution, while the complement of this fraction has no effect. Q-Q plots are then produced to estimate cross-phenotype enrichment. This analysis uses variable *p*-value thresholds for the SNPs association with a given trait as a function of their association with a second trait. If the Q-Q curves for phenotype one increasingly deflect left from the theoretical null line as the significance in phenotype two increases (*i.e.* become more stringent, reflecting stronger associations), this supports enrichment due to overlapping genomic signal and adds data to the visualization of the proportion of shared *vs.* unique polygenic signal provided by MiXeR. Existence of genetic overlap between pair of phenotypes warrants combining these phenotypes in conjFDR analysis.

### conjFDR details

Conjunctional false discovery rate analysis (conjFDR) estimates posterior probability that a given SNP is null for either phenotype or both phenotypes simultaneously when the p-values for both phenotypes are as small or smaller than the p-values observed in the original GWAS. To control for spurious enrichment in the conjFDR analyses, random pruning was averaged over 100 iterations (noticing that a higher number of iterations yielded identical results), and one SNP in each LD block (r2 > 0.1) was randomly selected from each iteration. SNPs within the major histocompatibility complex (MHC; genome build 19 location 25652429-33368333), the chromosomal region 8p23 (location 7200000-12500000), owing to their complex LD structures, to avoid biased FDR estimation.

### FUMA details

#### Genomic loci definition

Using FUMA, iIndependent SNPs were defined as those with r^2^ <0.6. Amongst them, lead SNPs were defined as showing r^2^ <0.1. Candidate SNPs were defined as all SNPs in LD r^2^ ≥0.6 with one of the independent significant SNPs in the locus and determined the borders of each loci. Loci less than 250 kb apart were merged, with the most significant SNP considered to be the lead SNP of the merged locus. Overlapping signals within complex LD regions were represented by single most significant SNPs. The 1000 Genomes Project European-ancestry haplotype panel provided reference LD maps (1000 Genomes Project Consortium et al., 2010).

#### Functional annotation

Positional gene sets (N =278) were chosen to complement evidence regarding SNPs and genes with more straightforward positional data. Among biological pathways, we selected canonical pathways (N =2,269), which are restricted to curated, core biological processes. We also examined tissue-specific gene expression (GTEx data v.8 <https://gtexportal.org/home/>) and developmental changes in gene expression in the brain (<https://www.brainspan.org/>).

We applied a novelty checking procedure using the list of candidate loci from conjFDR analysis, that is, using a series of genomic ranges jointly associated with a pair of phenotypes at conjFDR *p*<0.1 against SNPs listed in an in-house database that combines the latest version of GWAS catalog (<https://www.ebi.ac.uk/gwas/home>) and the latest GWASs that are published for these phenotypes. All genome-wide significant SNPs that were previously published and that lie within the candidate ranges of conjFDR make this range “not novel”. This procedure, although not necessarily exhaustive given the nature of data that are searched for, can thus be considered highly conservative regarding false novelty claims.

### Validation analyses details

For MiXeR analysis, AC-GSCAN data were used including the recently merged UK Biobank data (<https://genome.psych.umn.edu/index.php/GSCAN#GSCAN_dbGaP_.26_UK_Biobank>) but, for conjFDR, UK Biobank data could not be included since sample overlap may inflate conjFDR results. We had to perform these analyses with the discovery MOOD sample, since no other adequately-sized study for this trait was available at the time of study.

### Phenotype anayses details

Regression models were built as follows: (i) MOOD as the dependent variable and AC or AUD as the independent variable, (ii) AC or AUD as the dependent variable and MOOD as the independent variable and (iii) AUD as the dependent variable and both AC and MOOD as the independent variables, with an additional interaction term between AC and MOOD.
