## Supplementary Data for "Genetic overlap between mood instability and alcohol-related phenotypes suggests shared biological underpinnings"

### Validation with independent GWASs

#### UKB MOOD *vs.* alcohol-related phenotypes (MiXeR, condFDR)

Both the extent of polygenic overlap and genetic correlation were replicated for MOOD & GSCAN-AC (**Supplementary Figure 7, A, B**) with distinct log-likelihood optimum (**Supplementary Figure 7, C**) and positive AIC value when compared to the model with minimum overlap. However, as with MVP-AUD, MiXeR was not deemed reliable for MOOD & PGC-AUD, as indicated by oscillating log-likelihood profile (**Supplementary Figure 7, D**) producing large standard deviation of the amount of ‘causal’ variants (SD =1,600 for 900 variants), and negative AIC compared to both minimum and maximum overlap (noticing that there was also a lack of enrichment in the corresponding Q-Q plot, data not shown). Consequently, conjFDR for MOOD and PGC-AUD yielded only one joint significant locus (**Supplementary Figure 8**). Effect directions of lead SNPs identified in discovery conjFDR analyses demonstrated a good agreement between discovery and validation GWASs on alcohol-related phenotypes (**Supplementary Table 1**). For AC, 15 of 18 (83%) loci were concordant (*p* =0.0037), while for AUD 14 out of 20 (70%) were concordant (*p* =0.0576). Finally, in order to investigate whether MiXeR MOOD & AUD results were due to power issues or to the genetic architecture of AUD itself, we ran MiXeR between MOOD and a meta-analyzed summary statistics of AUD from PGC + MVP samples (**Supplementary Figure 9**). We showed that, while the statistical power was overall increased (**Supplementary Figure 9A**), MiXeR estimates of phenotype-specific and shared fraction of “causal” variants remained unstable, as indicated by large standard deviation, erratic log-likelihood plot (**Supplementary Figure 9B**). AIC values for both minimum and maximum possible overlap were marginally positive.

#### AC *vs.* AUD (MiXeR, conjFDR)

All MiXeR and genetic correlations analyses fully replicated the patterns of overlap between AC and AUD (**Supplementary Figure 10**). Interestingly, the GSCAN-AC & MVP-AUD MiXeR analysis showed a slightly different pattern, with a complete overlap between the traits and the highest correlation of the AC & AUD analyses (r_g_ =0.73). Importantly, conjFDR associations (**Supplementary Figure 11**) were replicated for four (AC) and five (AUD) loci, respectively, as reported in **Supplementary Table 1**.

### Supplementary Tables and Figures

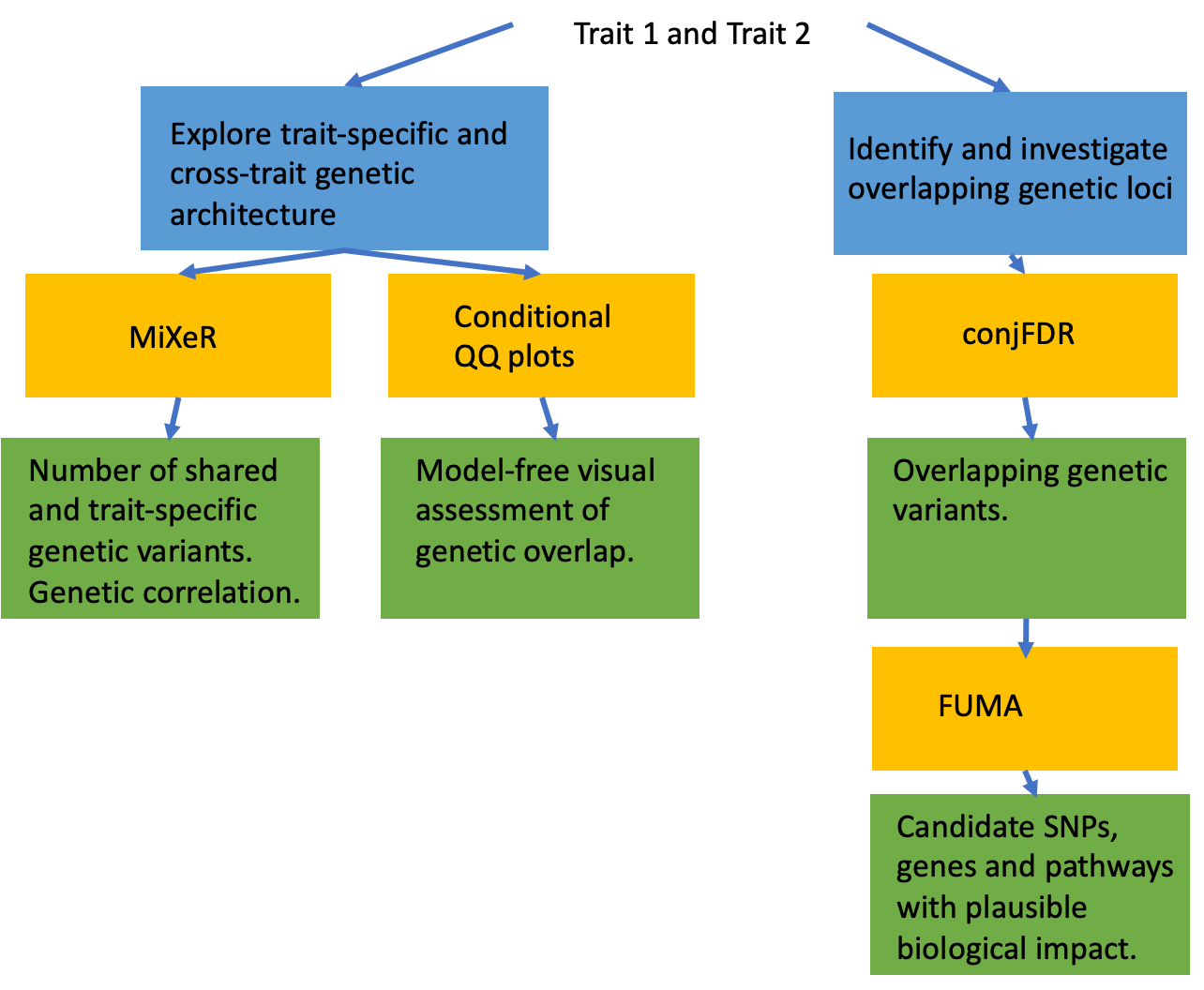
**Supplementary Figure 1:** overview of analyses and annotations

***Phenotype 1 and Phenotype 2***

| **MiXeR** | **conjFDR** |
| --- | --- |
| *Causal (i.e. beyond LD issues) SNPs ?* | |
| x |  |
| *Use of overlapping samples ?* | |
| x |  |
| *Individual loci for further functional characterization?* | |
|  | x |

FDR, false discovery rate; GWAS, genome-wide association study; FUMA, Functional Mapping and Annotation; SNP, single nucleotide polymorphism. Blue, scientific question; yellow, Analysis method/tool; green, output. The embedded table summarizes the comparative advantages of MiXeR *vs.* conjFDR.

| **Supplementary Figure2: log-likelihood plots from the discovery MiXeR analysis.** The number of causal variants is expressed in thousands. | | |
| --- | --- | --- |
| **A**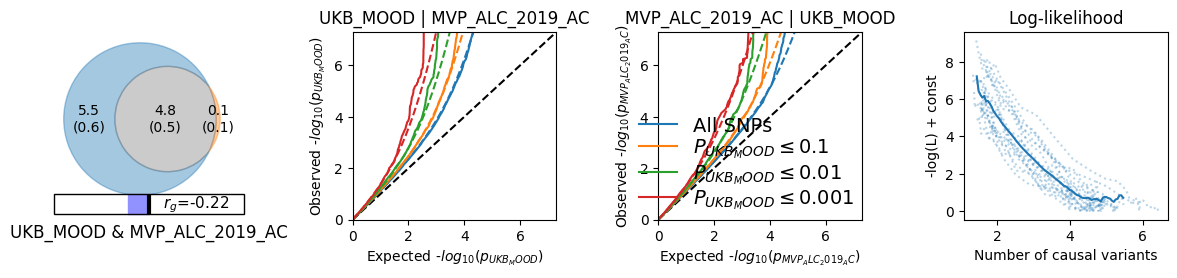 | **B** 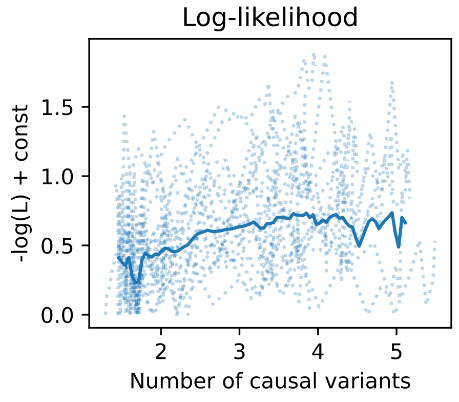 | **C** 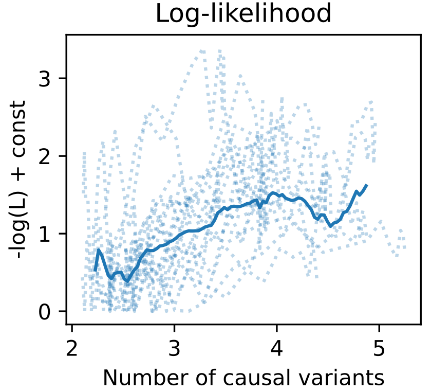 |
| MOOD, mood instability in the UK biobank; AC, alcohol consumption in the MVP sample; AUD, alcohol use disorder in the MVP sample. | | |

**Supplementary Figure 3:** Q-Q plots from MiXeR analyses showing enrichment for significant SNPs from trait one as the significance in the second trait increases.

A) AC|MOOD, MOOD|AC

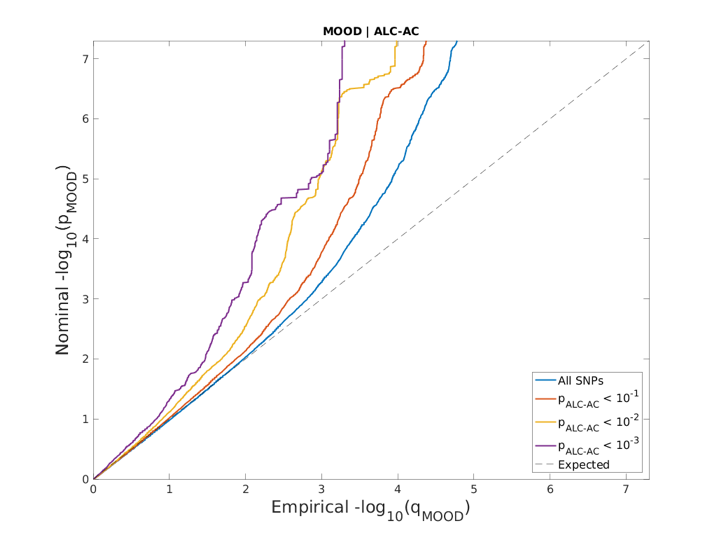

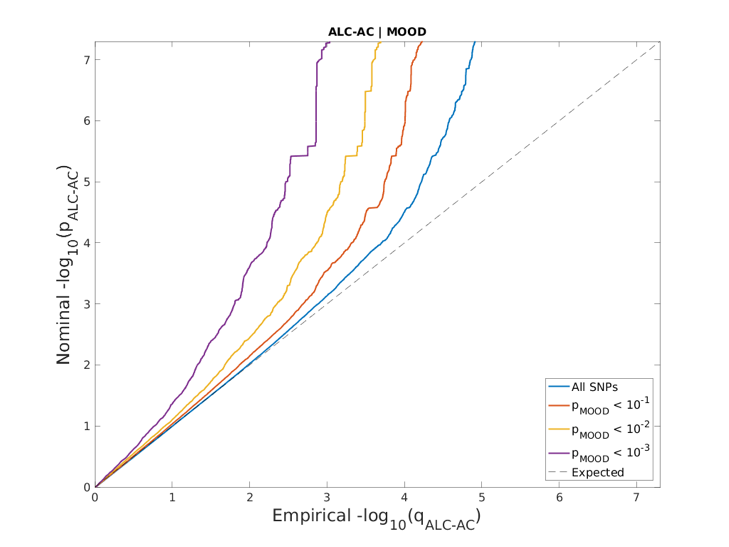

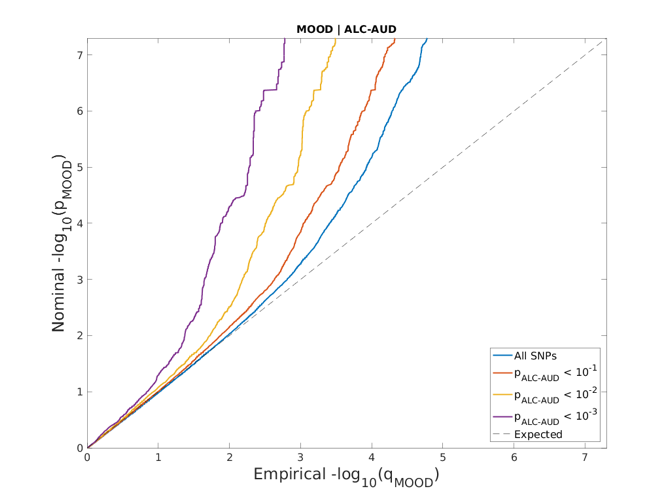

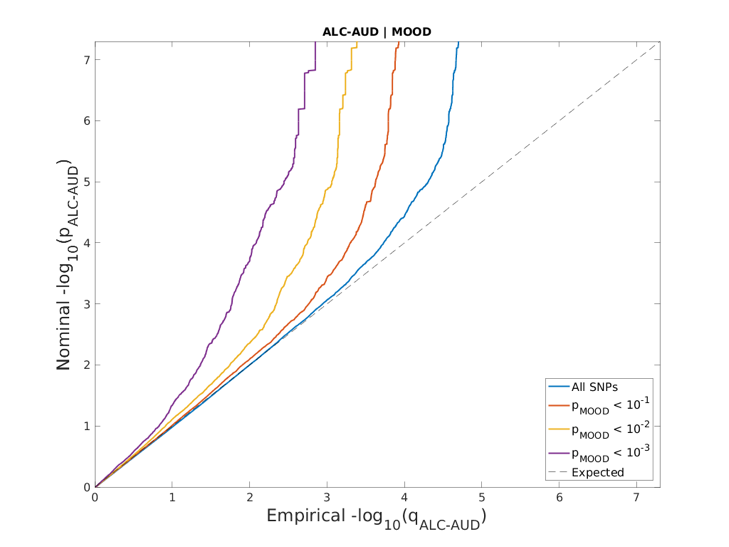
B) AUD|MOOD, MOOD|AUD

MOOD, mood instability in the UK biobank; AC, alcohol consumption in the MVP sample; AUD, alcohol use disorder in the MPV sample.

**Supplementary Figure 4:** signficantly enriched canonical pathways for (A) mood instability (MOOD) & alcohol consumption (AC) and (B) MOOD & alcohol use disorder (AUD). Gene sets were obtained by Functional Mapping and Annotation (FUMA) procedure based on the genes mapped from the discovery conjFDR analysis.

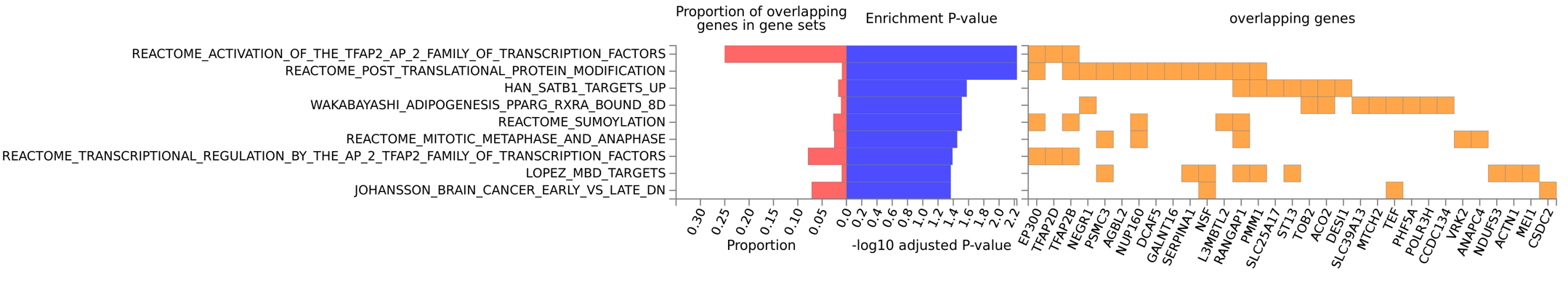
**A)**

**B)**

R

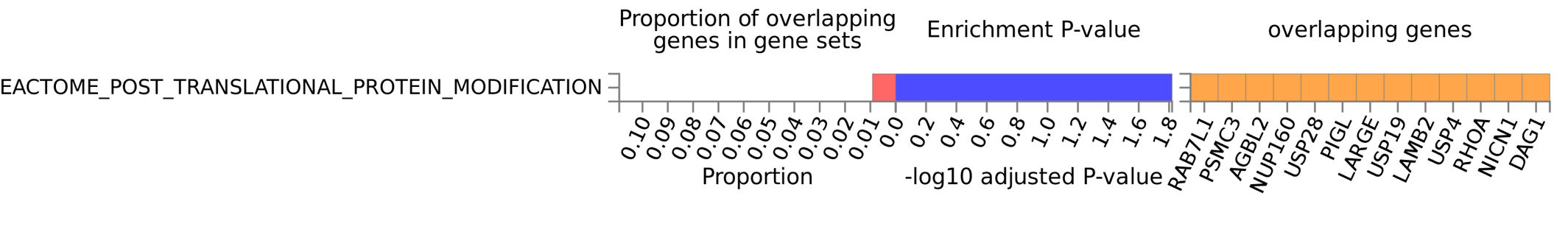

**Supplementary Figure 5: increasing *p*-values for tissue-specific differential gene expression (both sides)** for genes mapped after conjFDR for (A) mood instability and MVP-alcohol consumption and (B) mood instability and MVP-alcohol use disorder. Gene expression is obtained from GTEx V.8 (<https://gtexportal.org/home/>). Image was cropped, leaving some unwanted marks on panel B.

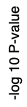

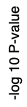

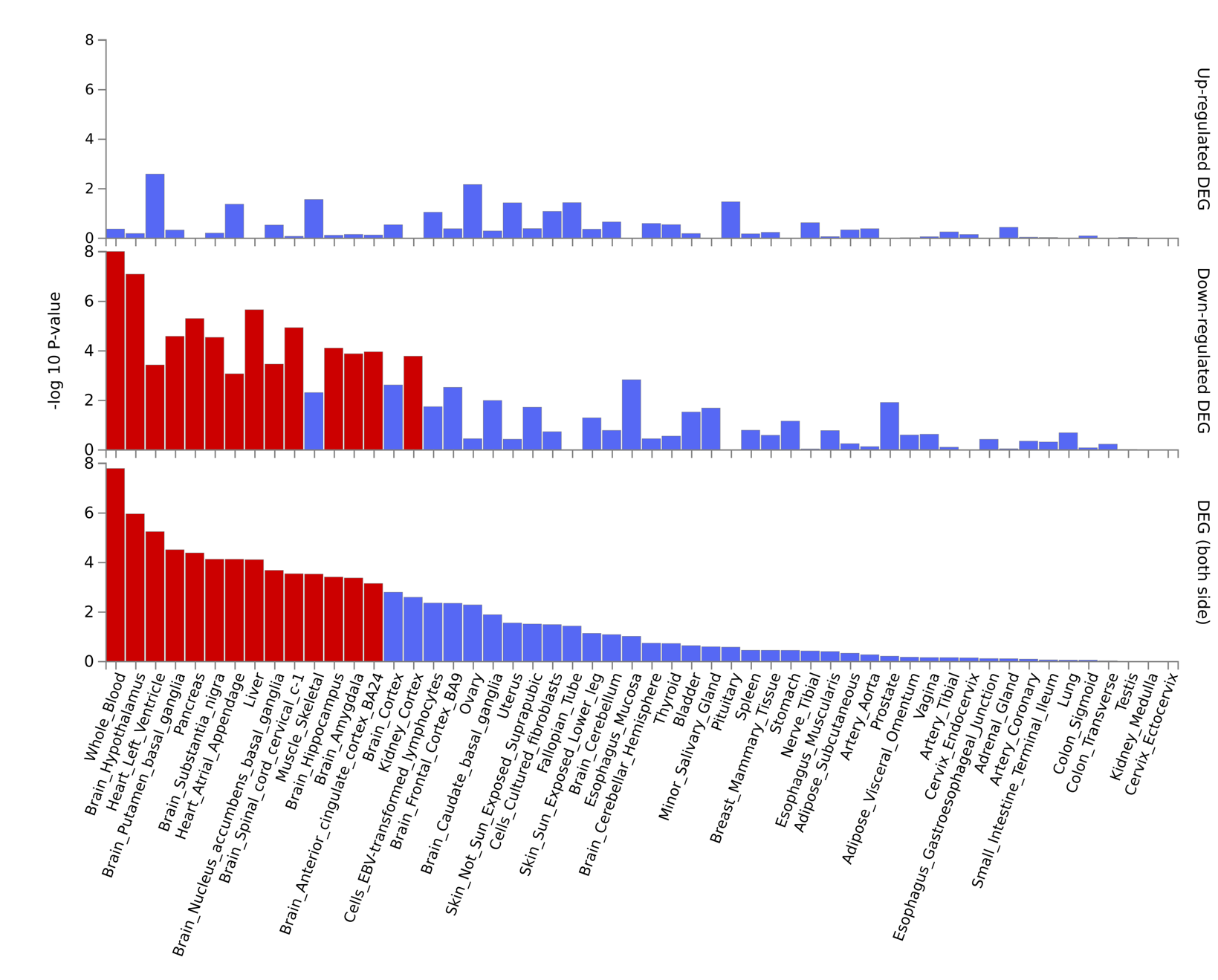

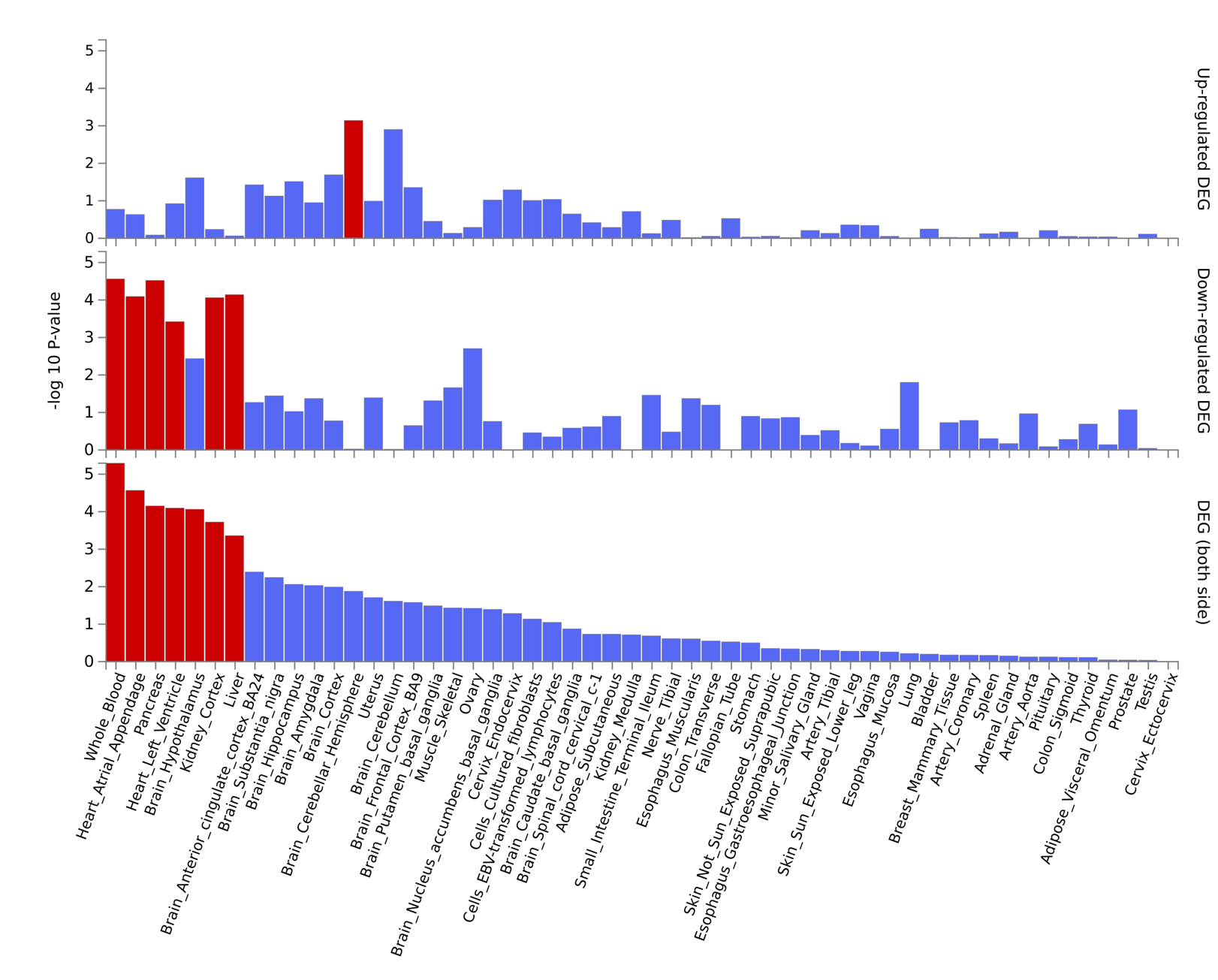

**A)	B)**

**Supplementary Figure 6: expression heatmaps** for genes mapped after conjFDR for (A) mood instability and MVP-alcohol consumption and (B) mood instability and MVP-alcohol use disorder. Gene expression is obtained from BrainSpan data (<https://www.brainspan.org/>) at various developmental stages during fetal life, infancy, adolescence and adulthood.

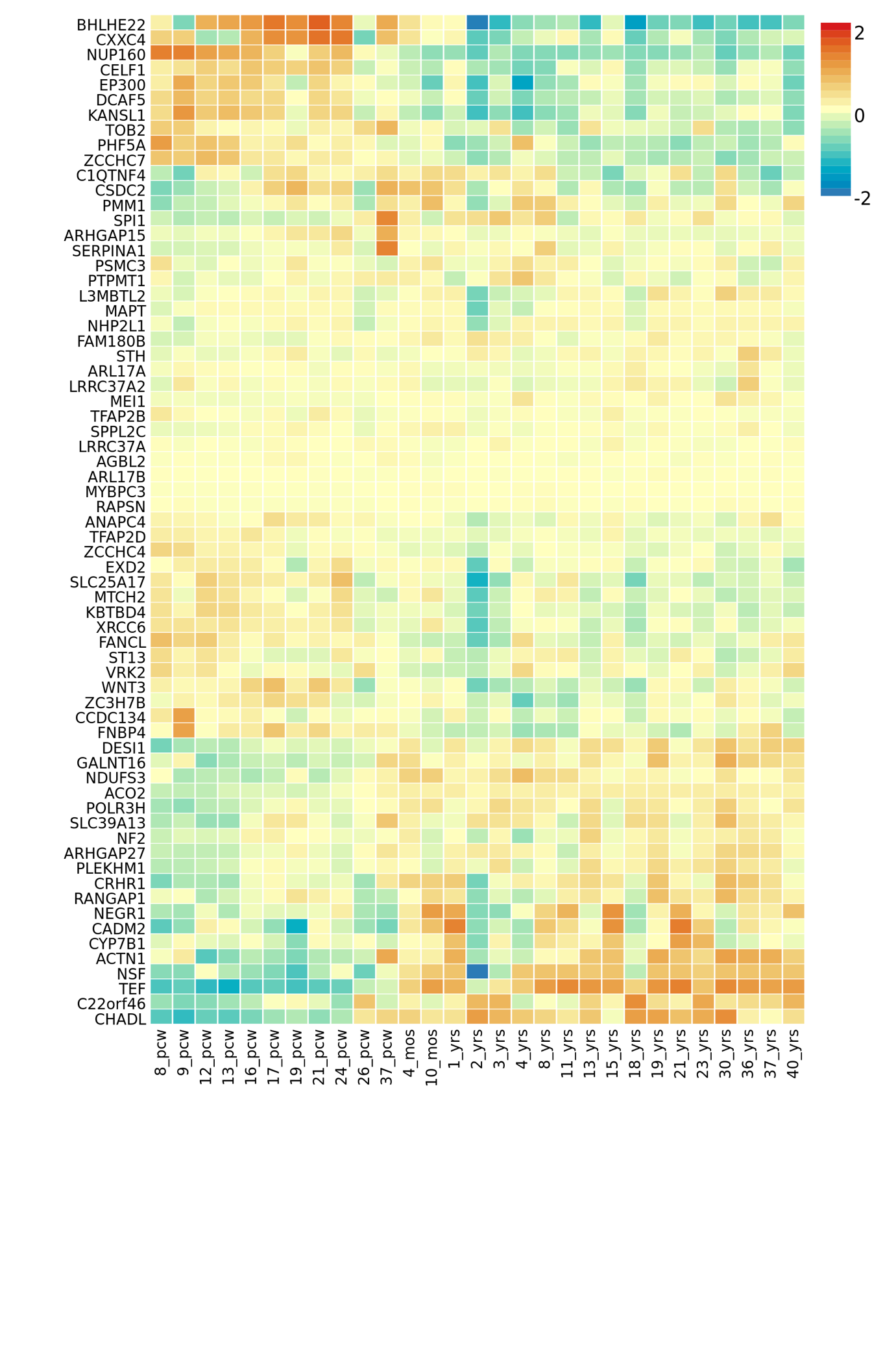

**A)**

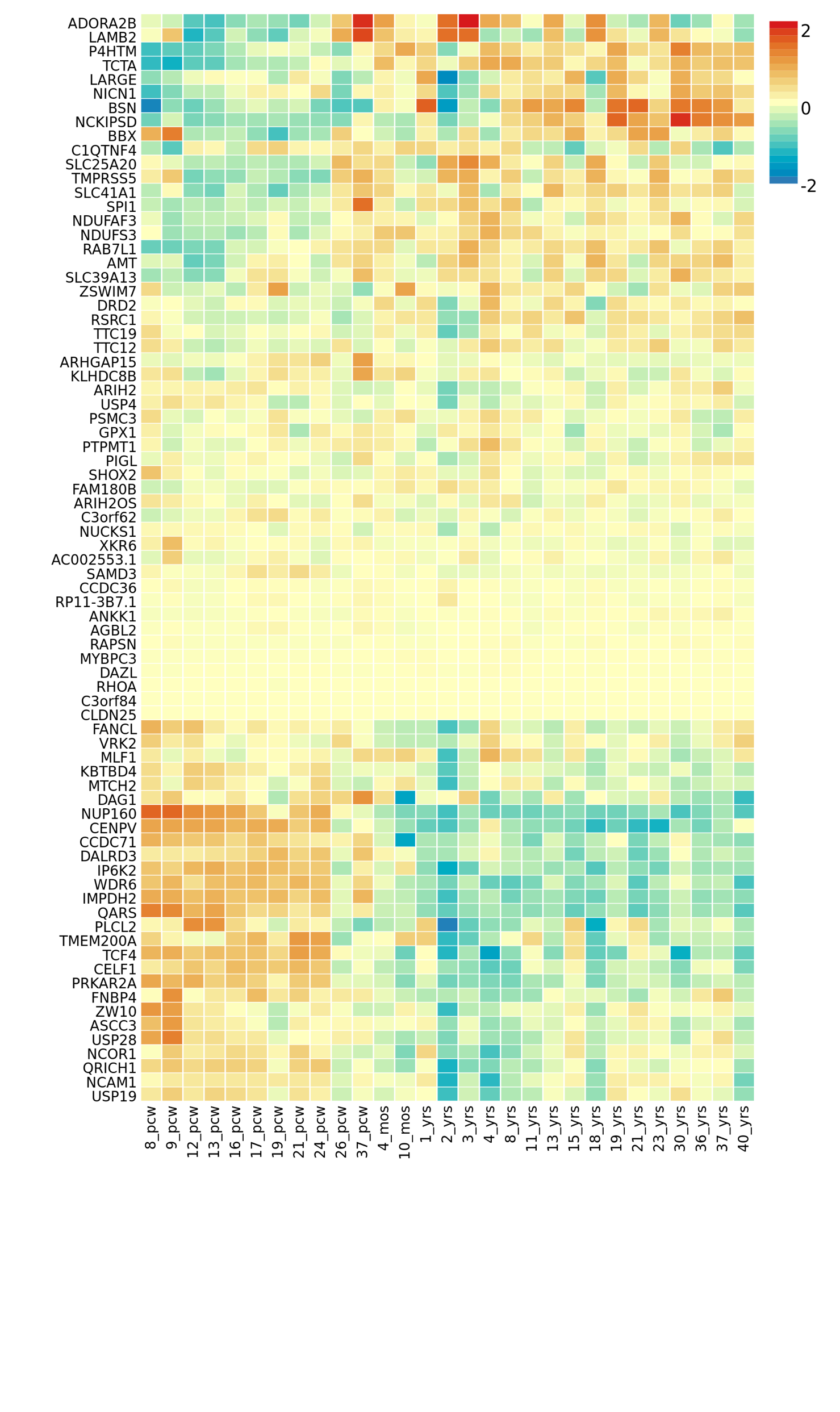
**B)**

| **Supplementary Figure 7: validation for mood instability and alcohol-related phenotypes.** Venn diagrams and Q-Q plots from MiXeR replication analyses showing polygenic overlap and enrichment for significant SNPs at decreasing thresholds. Analyses were performed using GWAS from UK biobank for mood instability (MOOD), GSCAN for alcohol consumption (AC_rep_) and PGC for alcohol use disorder (AUD_rep_). rg, genetic correlation. | |
| --- | --- |
| 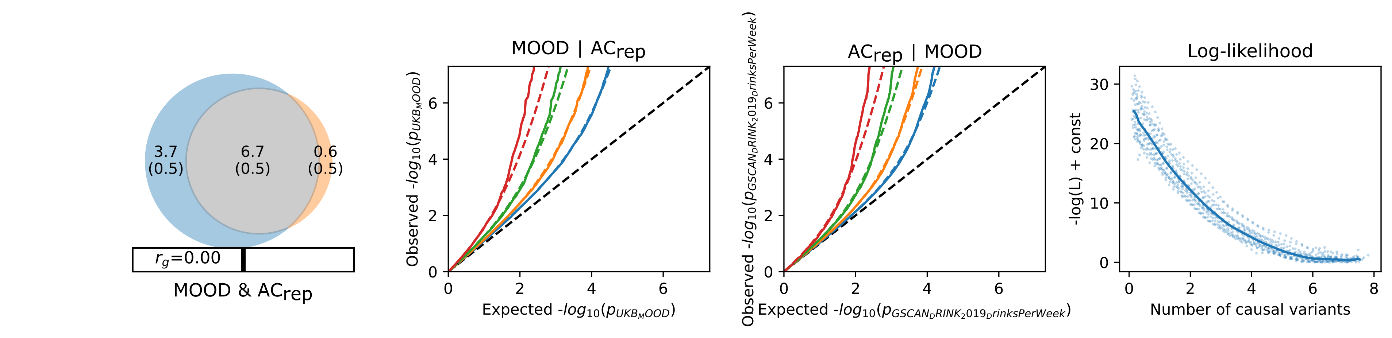**A** | 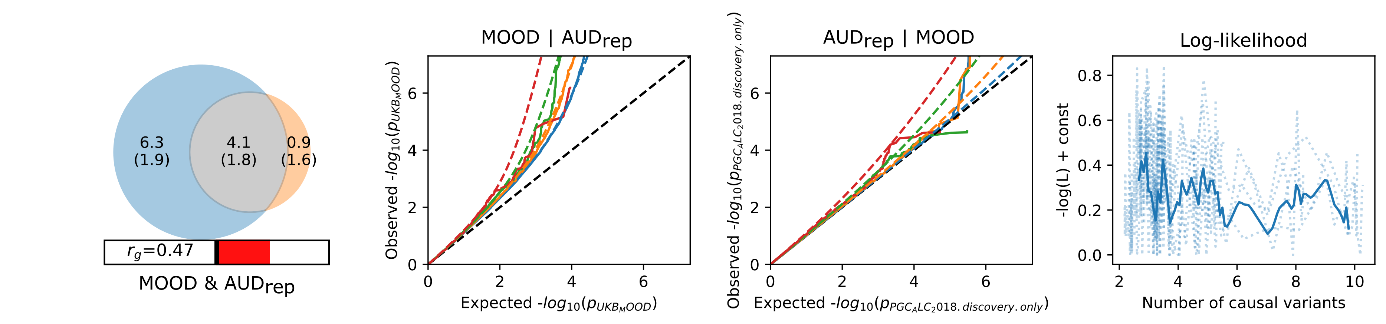**B** |
| **C** 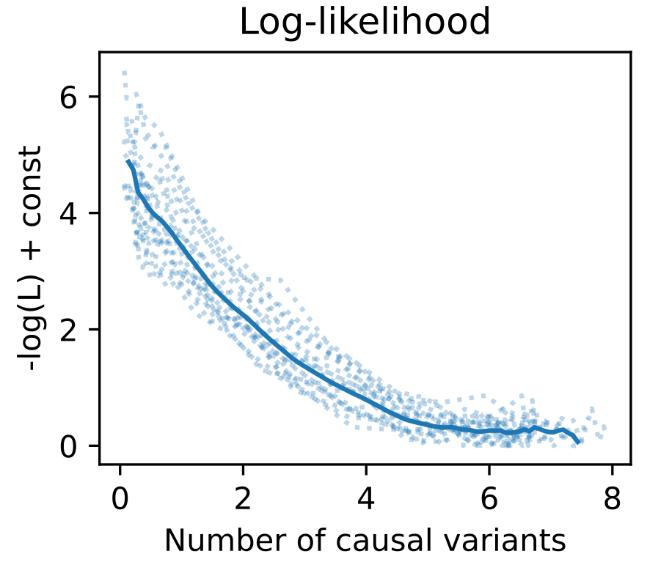 | **D** 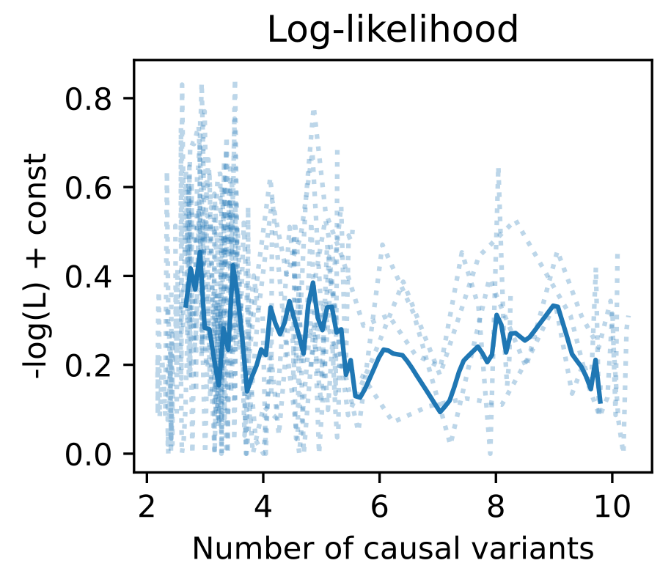 |

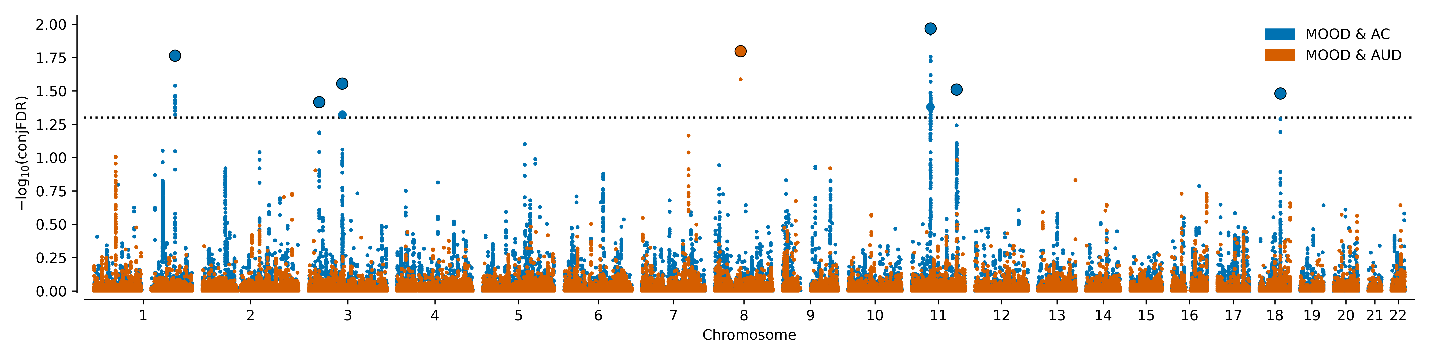
**Supplementary Figure 8: conjFDR validation for mood instability and alcohol-related phenotypes.** Manhattan plots for conjFDR between mood instability and GSCAN-alcohol consumption (MOOD & AC, in blue) and between mood instability and PGC-alcohol use disorder (MOOD & AUD, in brown).

**Supplementary Figure 9: Validation analyses for mood instability and alcohol use disorder using meta-analysis between the MVP and the PGC samples.** (A) Q-Q plots from MVP alone *vs.* MVP+PGC AUD summary statistics. (B) Venn diagrams and Q-Q plots from MiXeR replication analyses showing polygenic overlap and enrichment for significant SNPs at decreasing thresholds. Analyses were performed using GWAS from UK biobank for mood instability (MOOD). rg, genetic correlation.

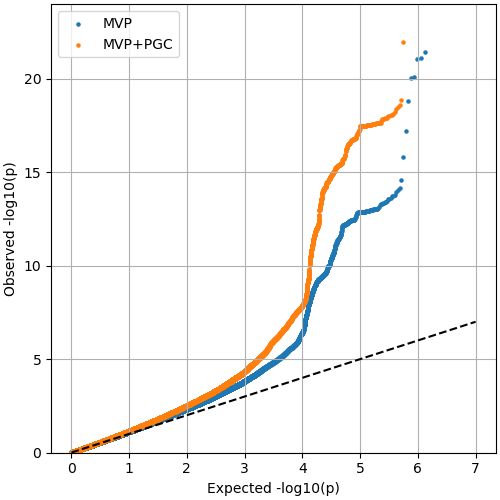
**(A)**

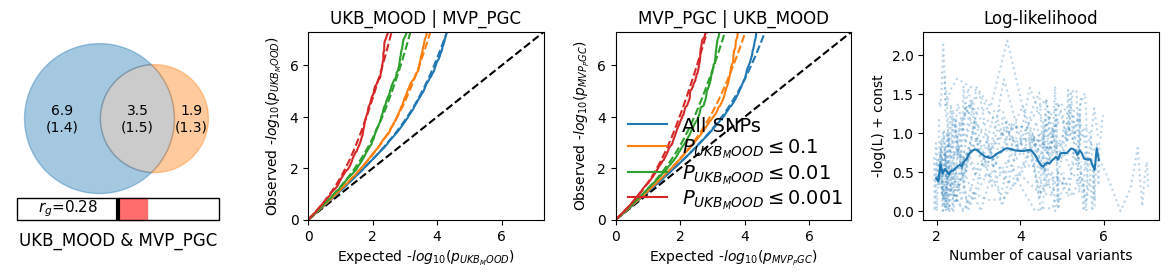
**(B)**

MOOD, mood instability in the UK biobank; MVP_PGC, meta-analysis of the alcohol use disorder in the Million Veteran Program + the Psychiatric Genomics Consortium samples.

| **Supplementary Figure 10: Validation for alcohol-related phenotypes.** Venn diagrams and log-likelihood plots from MiXeR quasi-replication showing polygenic overlap and enrichment for significant SNPs at decreasing thresholds. rg, genetic correlation. | |
| --- | --- |
| **A**  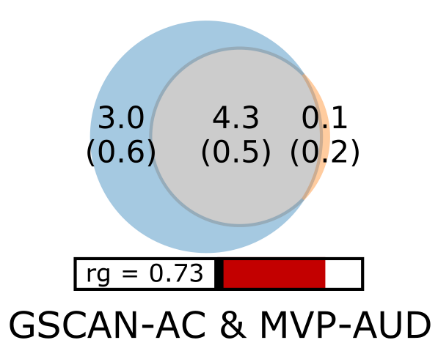 | **B**  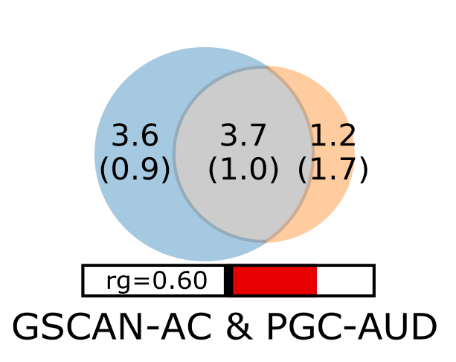 |
| **C**  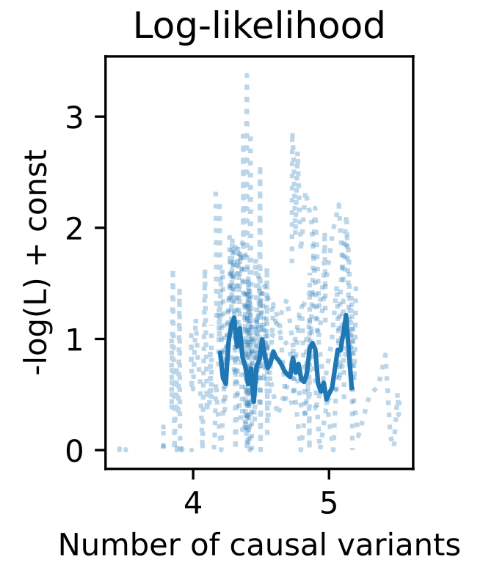 | **D**  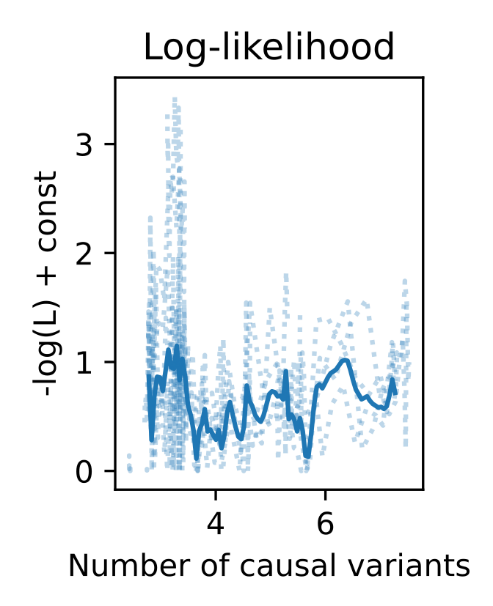 |
| MVP, Million Veteran Program; GSCAN, GWAS & Sequencing Consortium of Alcohol and Nicotine use; PGC, Psychiatric Genetics Consortium; AC, alcohol consumption; AUD, alcohol use disorder. | |

**Supplementary Figure 11:** Manhattan plots for conjFDR between GSCAN-alcohol consumption and MVP-alcohol use disorder (GSCAN-AC & MVP-AUD, in green) and between MVP-alcohol consumption and PGC-alcohol use disorder (MVP-AC & PGC-AUD, in yellow).

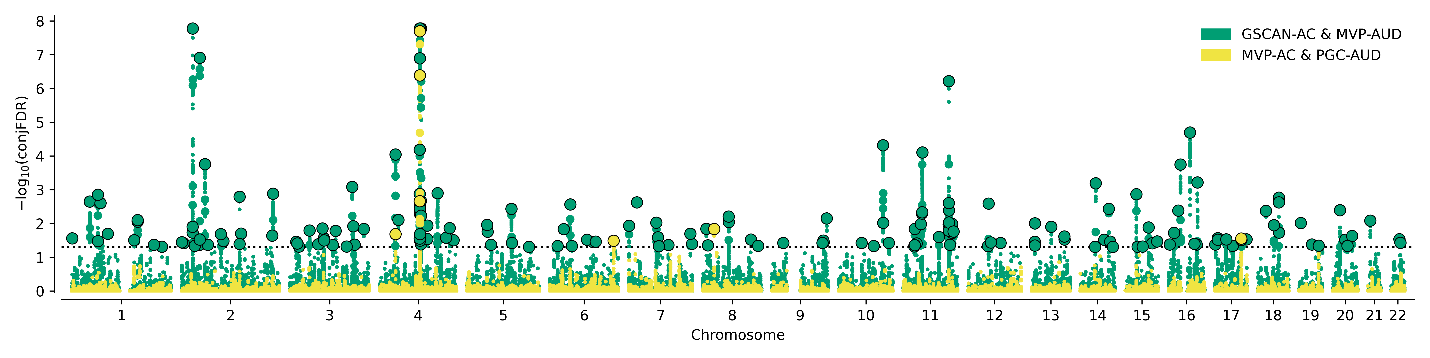

MVP, Million Veteran Program; GSCAN, GWAS & Sequencing Consortium of Alcohol and Nicotine use; PGC, Psychiatric Genetics Consortium. AC, alcohol consumption; AUD, alcohol use disorder.

**Supplementary Table 1:** significant lead SNPs from conjunctional false discovery rate (conjFDR) analysis and their functional annotation for (A) mood instability and alcohol consumption and (B) mood instability and alcohol use disorder. *P*-values and Z scores are rounded for five digits. Novel SNPs for GWASs about MOOD, AC and AUD published as of January, 28 2022 are written in bold.

A) **MOOD and AC**

| **CHR** | **Start_BP** | **End_BP** | **SNP** | **A1** | **A2** | **pval_MOOD** | **pval_AC** | **zscore_MOOD** | **zscore_AC** | **Nearest gene** | **Function** | **CADD** | **RDB** | **minChrState** |
| --- | --- | --- | --- | --- | --- | --- | --- | --- | --- | --- | --- | --- | --- | --- |
| **1** | **72628347** | **72949290** | **rs2568957** | **A** | **G** | **0.00004** | **0.00013** | **-4.1** | **3.8** | ***RPL31P12*** | **intergenic** | **2.758** | **7** | **14** |
| 2 | 22430795 | 22493637 | rs10865093 | C | T | 0.00046 | 0.00076 | 3.5 | -3.4 | *AC068490.2* | ncRNA_intronic | 8.817 | 7 | 9 |
| 2 | 57942987 | 58505679 | rs2312147 | T | C | <10^-5^ | <10^-5^ | -4.8 | -5.4 | *VRK2* | intronic | 3.184 | 6 | 7 |
| 2 | 144145478 | 144263280 | rs12691680 | T | C | <10^-5^ | 0.00047 | -5.5 | -3.5 | *ARHGAP15:AC096558.1:RP11-570L15.2* | ncRNA_intronic | 1.939 | 6 | 4 |
| 3 | 85370081 | 85387310 | rs818219 | C | T | 0.00001 | 0.00049 | -4.5 | 3.5 | *CADM2* | intronic | 1.378 | 6 | 5 |
| **4** | **25342606** | **25408838** | **rs34811474** | **A** | **G** | **0.00052** | **0.00083** | **-3.5** | **3.3** | ***ANAPC4*** | **exonic** | **23.7** | **6** | **4** |
| **4** | **105361545** | **105499837** | **rs17034592** | **C** | **T** | **0.00004** | **0.00062** | **-4.1** | **3.4** | ***AC004053.1*** | **ncRNA_intronic** | **7.711** | **7** | **5** |
| **6** | **50597378** | **50934086** | **rs4642472** | **T** | **C** | **0.00079** | **0.00004** | **-3.4** | **4.1** | ***FTH1P5*** | **intergenic** | **0.253** | **5** | **9** |
| **7** | **1221100** | **1228945** | **rs12533133** | **C** | **T** | **0.00074** | **0.00033** | **3.4** | **-3.6** | ***AC091729.9*** | **intergenic** | **4.537** | **2b** | **5** |
| 8 | 9177732 | 9231661 | rs11783641 | A | G | 0.00002 | 0.00004 | -4.3 | -4.1 | *RP11-115J16.1* | ncRNA_intronic | 4.894 | 4 | 5 |
| 8 | 65500967 | 65745896 | rs13260351 | T | C | 0.00001 | 0.00072 | -4.3 | 3.4 | *CYP7B1* | intronic | 0.189 | 6 | 15 |
| 9 | 37073902 | 37379492 | rs11544336 | G | A | 0.00098 | 0.00006 | -3.3 | 4 | *RP11-220I1.1* | ncRNA_exonic | 5.595 | NA | 4 |
| 11 | 47372377 | 47946836 | rs11039265 | A | C | 0.0001 | 0.00011 | -3.9 | -3.9 | *CELF1* | intronic | 0.059 | 5 | 4 |
| **14** | **69429386** | **69755470** | **rs8007859** | **G** | **T** | **0.00003** | **0.00003** | **-4.2** | **4.2** | ***EXD2*** | **exonic** | **9.825** | **5** | **4** |
| 14 | 94838142 | 94844947 | rs112635299 | T | G | 0.00002 | <10^-5^ | -4.3 | -4.6 | *SERPINA1* | intergenic | 0.272 | 5 | 5 |
| 17 | 43460181 | 44865603 | rs199518 | A | C | <10^-5^ | 0.00005 | 6.5 | -4.1 | *WNT3* | intronic | 5.631 | 5 | 5 |
| **22** | **30040197** | **30095706** | **rs2857651** | **C** | **T** | **0.00069** | **0.00006** | **3.4** | **-4** | ***NF2*** | **intronic** | **11.18** | **7** | **4** |
| 22 | 41215672 | 42216326 | rs75723348 | G | T | <10^-5^ | <10^-5^ | 5.8 | -4.8 | *Y_RNA* | intergenic | 5 | 6 | 5 |

CHR; chromosome, BP, base pair; SNP, single nucleotide polymorphism; A1, alternate allele; A2, reference allele; MOOD, mood instability; AC, alcohol consumption; CADD, Combined Annotation Dependent Depletion; RDB, RegulomeDB; minChrState, chromatin state.

B) **MOOD and AUD**

| **CHR** | **Start_BP** | **End_BP** | **SNP** | **A1** | **A2** | **pval_MOOD** | **pval_AUD** | **zscore_MOOD** | **zscore_AUD** | **nearestGene** | **func** | **CADD** | **RDB** | **minChrState** |
| --- | --- | --- | --- | --- | --- | --- | --- | --- | --- | --- | --- | --- | --- | --- |
| 1 | 73736562 | 74108328 | rs7540134 | T | C | 0.00111 | 0.00051 | 3.3 | 3.5 | RP4-788P17.1 | intergenic | 7.553 | 5 | 5 |
| 1 | 205674897 | 205765040 | rs144482900 | GA | G | 0.00077 | 0.00064 | -3.4 | -3.4 | NUCKS1:AC119673.1 | exonic | 34 | NA | 4 |
| 2 | 57942987 | 58505679 | rs2312147 | T | C | <10^-5^ | 0.00001 | -4.8 | -4.5 | VRK2 | intronic | 3.184 | 6 | 7 |
| 2 | 144145478 | 144263280 | rs4273169 | A | G | <10^-5^ | 0.00006 | -5.5 | -4 | ARHGAP15:AC096558.1:RP11-570L15.2 | ncRNA_intronic | 17.32 | 5 | 2 |
| 3 | 16716571 | 17119238 | rs9882937 | T | C | 0.00031 | 0.00011 | 3.6 | 3.9 | PLCL2 | intronic | 6.327 | 5 | 5 |
| **3** | **48724599** | **49650935** | **rs11130187** | **T** | **C** | **0.00002** | **0.00002** | **4.2** | **4.2** | **C3orf62** | **UTR3** | **0.873** | **7** | **4** |
| **3** | **107237696** | **107379837** | **rs600011** | **C** | **A** | **<10^-5^** | **0.00027** | **5.8** | **-3.6** | **BBX** | **intronic** | **2.719** | **6** | **5** |
| 3 | 157829953 | 158284861 | rs827802 | A | C | 0.00034 | 0.0001 | -3.6 | -3.9 | RSRC1 | intronic | 7.349 | NA | 5 |
| **6** | **100953047** | **101505612** | **rs9390701** | **A** | **G** | **0.0002** | **0.00041** | **3.7** | **3.5** | **RP3-467N11.2** | **intergenic** | **7.053** | **6** | **5** |
| **6** | **130544509** | **130768029** | **rs7773962** | **G** | **A** | **0.00009** | **0.00054** | **-3.9** | **-3.5** | **SAMD3** | **intronic** | **2.832** | **5** | **1** |
| **7** | **1221100** | **1228945** | **rs34527042** | **C** | **T** | **0.00057** | **0.00039** | **3.4** | **-3.5** | **AC091729.9** | **intergenic** | **10.78** | **6** | **5** |
| 8 | 9177732 | 9239958 | rs66541322 | T | C | 0.00011 | 0.00009 | -3.9 | -3.9 | RP11-115J16.1:RP11-115J16.3 | ncRNA_intronic | 3.21 | 7 | 5 |
| **8** | **10876819** | **10955220** | **rs7833741** | **C** | **A** | **0.00001** | **0.00012** | **4.4** | **3.8** | **XKR6** | **intronic** | **1.985** | **7** | **5** |
| 11 | 47372377 | 47946836 | rs11039216 | C | T | <10^-5^ | <10^-5^ | -5.3 | -4.8 | RP11-750H9.5 | ncRNA_intronic | 0.987 | 6 | 5 |
| 11 | 112826867 | 112938783 | rs1940701 | C | T | 0.00016 | 0.00032 | 3.8 | 3.6 | NCAM1 | intronic | 5.209 | NA | 5 |
| 11 | 113199146 | 113692660 | rs7933981 | A | G | <10^-5^ | <10^-5^ | -5.1 | -6 | DRD2 | intergenic | 17.19 | 6 | 5 |
| **13** | **68141716** | **68194993** | **rs9541161** | **T** | **C** | **0.00085** | **0.00053** | **-3.3** | **3.5** | **LINC00364** | **intergenic** | **3.989** | **7** | **9** |
| **17** | **15873275** | **16263682** | **rs3785620** | **C** | **A** | **0.00006** | **0.00064** | **4** | **-3.4** | **PIGL** | **intronic** | **0.454** | **6** | **4** |
| 18 | 52876850 | 53389061 | rs2958171 | T | C | <10^-5^ | 0.00006 | 5.2 | -4 | TCF4 | intronic | 9.216 | 4 | 1 |
| **22** | **34227271** | **34269594** | **rs2277840** | **G** | **A** | **0.00005** | **0.00031** | **4.1** | **3.6** | **LARGE** | **intronic** | **3.942** | **7** | **5** |

CHR; chromosome; BP, base pair; SNP, single nucleotide polymorphism; A1, alternate allele; A2, reference allele; MOOD, mood instability; AUD, alcohol use disorder; CADD, Combined Annotation Dependent Depletion; RDB, RegulomeDB; minChrState, chromatin state.

**Supplementary Table 2:** overview of polygenic overlap and genetic correlation obtained by MiXeR in each discovery and validation analysis. Note that the degree of polygenic overlap is based on 90% of the joint genetic signal for the two phenotypes considered.

| **Phenotype 1** | **Phenotype 2** | **Sample 1** | **Sample 2** | **% shared polygenic overlap for phenotype 1** | **% shared polygenic overlap for phenotype 2** | **Genetic correlation (r_g_)** |
| --- | --- | --- | --- | --- | --- | --- |
| *Discovery analyses* | | | | | | |
| MOOD | AC | UKB | MVP | 47 | 98 | -0.22 |
|  | AUD |  |  | 20 | 49 | 0.23 |
| AC | AUD |  |  | 51 | 58 | 0.52 |
| *Validation* | | | | | | |
| MOOD | AC | UKB | GSCAN | 74 | 92 | 0 |
|  | AUD | UKB |  | 39 | 82 | 0.47 |
| AC | AUD | GSCAN | MVP | 59 | 98 | 0.73 |
|  |  | MVP |  | 51 | 58 | 0.52 |
|  |  | GSCAN | PGC | 51 | 76 | 0.6 |
|  |  | GSCAN (without UKB) | PGC | 45 | 61 | 0.52 |
| AC | MDD | MVP  MVP_PGC | PGC | 98 | 35 | -0.2 |
| AUD |  |  |  | 65 | 25 | 0.4 |

MOOD, mood instability; AC, alcohol consumption; AUD, alcohol use disorder; UKB, UK Biobank; MVP, Million Veteran Program; GSCAN, GWAS & Sequencing Consortium of Alcohol and Nicotine use; PGC, Psychiatric Genetics Consortium; MDD, major depressive disorder; MVP_PGC, meta-analysis of the alcohol use disorder in the Million Veteran Program + the Psychiatric Genomics Consortium samples**.**

**Supplementary Table 3:** Lead SNPs with validation conjFDR *p* <0.05 for alcohol-related phenotypes.

| **SNP** | **CHR** | **BP** | **A1** | **A2** | **conjFDR**  **discovery** | **Z or BETA**  **discovery** | **conjFDR**  **validation** | **Z or BETA**  **validation** |
| --- | --- | --- | --- | --- | --- | --- | --- | --- |
| *MVP-AC & GSCAN-AC* | | | | | | | | |
| **rs2312147** | 2 | 58222928 | C | T | 7.27E-09 | 0.03166 | 0.00414 | 0.00883 |
| **rs13411140*** | 2 | 144215811 | C | T | 0.0002872 | 0.02014 | 0.00435 | 0.00885 |
| **rs818219** | 3 | 85374589 | C | T | 0.0002052 | 0.01989 | 0.000324 | 0.0108 |
| **rs112635299** | 14 | 94838142 | G | T | 8.06E-07 | 0.1043 | 0.000492 | 0.0405 |
| **rs11039255*** | 11 | 47495746 | G | T | 5.18E-05 | 0.02297 | 0.00102 | 0.0103 |
| *MVP-AUD & PGC-AUD* | | | | | | | | |
| **rs4273169** | 2 | 144231309 | A | G | 2.63E-05 | -4.204 | 0.00957 | -2.591 |
| **rs1940701** | 11 | 112869404 | C | T | 0.0001677 | 3.763 | 0.03791 | 2.076 |
| **rs7933981** | 11 | 113438068 | A | G | 3.3E-10 | -6.284 | 0.007097 | -2.692 |
| **rs2958171** | 18 | 53072832 | C | T | 2.85E-05 | 4.185 | 0.03127 | -2.154 |

SNP, single nucleotide polymorphism; CHR, chromosome; BP, position in base pairs; A1, alternate allele; A2, reference allele; MVP, Million Veteran Program; GSCAN, GWAS & Sequencing Consortium of Alcohol and Nicotine use; AC, alcohol consumption; AUD, alcohol use disorder; PGC, Psychiatric Genetics Consortium.

| **SNP** | **CHR** | **Phenotype with GWAS association ouside of MOOD, AC and AUD** | ***p-*value** | **Alcohol-related phenotype** | **PubMed ID** |
| --- | --- | --- | --- | --- | --- |
| rs600011 | 3 | Depressed affect | 2E-10 | AUD | 29942085 |
| rs600011 | 3 | Experiencing mood swings | 5E-09 | AUD | 29500382 |
| rs34811474 | 4 | Leisure sedentary behaviour (television watching) | 2E-09 | AC | 32317632 |
| rs34811474 | 4 | Snoring | 1E-08 | AC | 32060260 |
| rs34811474 | 4 | Snoring | 1E-10 | AC | 32060260 |
| rs34811474 | 4 | Predicted visceral adipose tissue | 7E-11 | AC | 31501611 |
| rs34811474 | 4 | Multisite chronic pain | 3E-11 | AC | 31194737 |
| rs34811474 | 4 | Snoring | 4E-11 | AC | 30804565 |
| rs34811474 | 4 | Osteoarthritis | 2E-09 | AC | 30664745 |
| rs34811474 | 4 | Heel bone mineral density | 3E-09 | AC | 30598549 |
| rs34811474 | 4 | White blood cell count | 2E-08 | AC | 30595370 |
| rs34811474 | 4 | Lung function (FVC) | 4E-11 | AC | 30595370 |
| rs34811474 | 4 | Height | 6E-18 | AC | 30595370 |
| rs34811474 | 4 | Heel bone mineral density | 2E-08 | AC | 30595370 |
| rs34811474 | 4 | Body mass index | 1E-32 | AC | 30595370 |
| rs34811474 | 4 | Male-pattern baldness | 4E-09 | AC | 30573740 |
| rs34811474 | 4 | Body mass index | 9E-38 | AC | 30239722 |
| rs34811474 | 4 | Body mass index | 9E-38 | AC | 30239722 |
| rs34811474 | 4 | Body mass index | 9E-38 | AC | 30239722 |
| rs34811474 | 4 | Body mass index | 2E-09 | AC | 30108127 |
| rs34811474 | 4 | Self-reported math ability (MTAG) | 7E-09 | AC | 30038396 |
| rs34811474 | 4 | Highest math class taken (MTAG) | 1E-13 | AC | 30038396 |
| rs34811474 | 4 | Educational attainment (years of education) | 3E-09 | AC | 30038396 |
| rs34811474 | 4 | Educational attainment (MTAG) | 7E-15 | AC | 30038396 |
| rs34811474 | 4 | Cognitive performance (MTAG) | 2E-18 | AC | 30038396 |
| rs34811474 | 4 | Cognitive performance | 1E-15 | AC | 30038396 |
| rs34811474 | 4 | Intelligence | 7E-16 | AC | 29942086 |
| rs34811474 | 4 | General cognitive ability | 7E-09 | AC | 29844566 |
| rs34811474 | 4 | Intelligence (MTAG) | 2E-10 | AC | 29326435 |
| rs34811474 | 4 | Body mass index | 3E-30 | AC | 29273807 |

**Supplementary Table 4: associations of novel SNPs with GWASs, other than MOOD, AC or AUD** according to GWAScatalog.

SNP, single nucleotide polymorphism; CHR, chromosome; MOOD, mood instability; AC, alcohol consumption; AUD, alcohol use disorder.
